# Predictive modeling of secondary pulmonary hypertension in left ventricular diastolic dysfunction

**DOI:** 10.1101/2020.04.23.20073601

**Authors:** Karlyn K. Harrod, Jeffrey L. Rogers, Jeffrey A. Feinstein, Alison L. Marsden, Daniele E. Schiavazzi

## Abstract

Diastolic dysfunction is a common pathology occurring in about one third of patients affected by heart failure. This condition is not associated with a marked decrease in cardiac output or systemic pressure and therefore is more difficult to diagnose than its systolic counterpart. Compromised relaxation or increased stiffness of the left ventricle with or without mitral valve stenosis induces an increase in the upstream pulmonary pressures, and is classified as secondary or group II (2018 Nice classification) pulmonary hypertension. This may result in an increase in the right ventricular afterload leading to right ventricular failure. Elevated pulmonary pressures are therefore an important clinical indicator of diastolic heart failure (sometimes referred to as *heart failure with preserved ejection fraction*, HFpEF), showing significant correlation with associated mortality. Accurate measurements of this quantity, however, are typically obtained through invasive catheterization, and after the onset of symptoms. In this study, we use the hemodynamic consistency of a differential-algebraic circulation model to predict pulmonary pressures in adult patients from other, possibly non-invasive, clinical data. We investigate several aspects of the problem, including the ability of model outputs to represent a sufficiently wide pathologic spectrum, identifiability of its parameters, to the accuracy of the predicted pulmonary pressures. We also find that a classifier using the assimilated model parameters as features is free from the problem of missing data and is able to detect pulmonary hypertension with sufficiently high accuracy. For a cohort of 82 patients suffering from various degrees of heart failure severity we show that systolic, diastolic and wedge pulmonary pressures can be estimated on average within 8, 6 and 6 mmHg, respectively. We also show that, in general, increased data availability leads to improved predictions.

## 1 Introduction

Diastolic heart failure (sometimes referred to as *heart failure with preserved ejection fraction* or HFpEF, see Table 1) is a serious, often fatal, cardiovascular pathology. Even though recent reviews [1] report how this pathology is often associated with a number of co-morbidities, making the selection of homogeneous treatment groups difficult, it is commonly characterized by elevated left ventricle filling pressures, but normal systemic circulatory indicators, such as left ventricular ejection fraction, cardiac output and mean arterial pressure [2]. In 2013, heart failure was mentioned in 1 of every 9 death certificates in the United States, and was the underlying condition in roughly 20% of these cases. The number of deaths attributable to heart failure was approximately as high in 1995 as it was in 2013, with hospital discharges remaining stable from 2000 to 2010 [3]. It is also estimated that about one-third of the patients with congestive heart failure (CHF) have a normal left ventricular ejection fraction (LVEF) [4].

**Table 1:** List of acronyms.

| Acronym | Description | Acronym | Description |
| --- | --- | --- | --- |
| CHF | Congestive heart failure | PH | Pulmonary hypertension |
| CVP | Central venous pressure | PVR | Pumonary vascular resistance |
| CO | Cardiac output | NM | Nelder-Mead |
| DBP | Diastolic blood pressure | MAP | Maximum a posteriori |
| dPAP | Diastolic pulmonary arterial pressure | MCMC | Markov chain Monte Carlo |
| EHR | Electronic health record | MPAP | Mean pulmonary arterial pressure |
| FIM | Fisher information matrix | RAP | Mean right atrial pressure |
| HFpEF | Heart failure with preserved ejection fraction | REDCap | Research electronic data capture |
| HR | Heart rate | RC | Resistance-capacitance |
| KL | Kullback-Leibler | RCR | Resistance-capacitance-resistance |
| LPN | Lumper parameter network | RLC | Resistance-inductance-capacitance |
| LVEDV | Left ventricular end diastolic volume | RVEDP | Right ventricular end diastolic pressure |
| LVEF | Left ventricular ejection fraction | RVEF | Right ventricular ejection fraction |
| OOP | Object oriented programming | SBP | Systolic blood pressure |
| PAP | Pulmonary artery pressure | sPAP | Systolic pulmonary arterial pressure |
| PCW | Pulmonary capillary wedge pressure | SVR | Systemic vascular resistance |

It has been observed how pulmonary hypertension (PH) is highly prevalent and often severe in HFpEF and that both pulmonary venous and arterial hypertension contribute to the severity of HFpEF with a marked correlation between systolic pulmonary arterial pressure (sPAP) and mortality [5, 1]. A recent paper reviewing the current understanding of etiology and treatment of HFpEF, reports how multiple nondiastolic abnormalities contribute to the syndrome of HFpEF, including LV systolic dysfunction, pulmonary hypertension, RV and LA dysfunction, vascular stiffening, ventricular interdependence and coronary microcirculation dysfunction [1]. Additionally, homogeous treatment HFpEF is made difficult by multiple phenotypes induced by obesity, ischemia and cardiometabolic abnormalities. This is consistent with the findings in [6] where a machine learning-based approach is used to cluster patients with HFpEF in three phenogroups with common characteristics, providing a way to go beyond a homogeneous treatment of HFpEF that has so far produced unsatisfactory results. Even though these studies highlight the current challenges in the treatment of HFpEF, they both seem to agree on the strong association of PH and HFpEF. In this regard [1] mentions how “PH is extremely common in HFpEF, seen in roughly 80% of patients, and mortality is increased in this cohort”, whereas in [6], elevated PH was one of the main criteria for patient recruitment.

While non invasive echocardiography and machine learning may be useful for phenotyping classification and treatment selection [1], early diagnosis of HFpEF relies on invasive pressure acquisition through right heart catheterization, often performed following the manifestation of symptoms [7, 8]. It is therefore evident how methods enabling an accurate indirect estimation of pulmonary pressures using minimally invasive clinical data would be extremely beneficial for early diagnosis of HFpEF, in a way that could trigger lifestyle changes that will, in turn, prevent other co-morbidities to develop.

In this study, we investigate how the physics-based consistency of a lumped parameter hemodynamic model containing three compartments, i.e., a four-chamber heart, systemic and pulmonary circulation compartments, may be used to monitor PH in patients from non-invasive and uncertain clinical measurements. The development of computer models to study hemodynamics in humans started in the 1960s and 1980s [9, 10, 11, 12], with application in pediatrics developed in the 2000s for single-ventricle congenital heart disease [13], Norwood physiology [14], and systemic-to-pulmonary artery shunts [15].

Approaches for automatic parameter estimation date back to the late 1970s [16, 17], ranging from two-stage Prony-Marquardt optimization [18], to adaptive control systems for left ventricular bypass assist devices [19, 20], to Kalman filters [21, 22] and to recursive least squares [23]. An iterative, proportional gain-based identification method is presented in [24], with application to coronary artery disease in [25]. Other studies include estimation of three-element Windkessel boundary conditions [26] and left ventricular viscoelasticity [27, 28]. More recently, examples of automatic parameter tuning in lumped circulatory models have included the physiology of children with congenital heart disease undergoing the first stage (i.e., Norwood) of single ventricle palliation surgery [29], construction of optimally trained patient-specific models for coronary artery disease [30] and predicting time evolution of ventricular dilation and thickening [31]. A study using lumped parameter models in diastolic heart failure is finally discussed in [32], while parameter identification for a mice model with chronic hypoxia and drug-induced pulmonary hypertension is proposed in [33].

Circuit models in hemodynamics typically contain tens of parameters which need to be trained from clinical records collected at multiple visits, which include a variable but typically sparse number of clinical measurements. This aggravates the ill-conditioning of the inverse problem, where model outputs do not change in response to perturbations along a number of unidentifiable linear combinations of parameters. In these circumstances, optimization may not be successful in identifying global optima and sequential Monte Carlo techniques [34] may underperform in practice, as data typically represent extremes (maxima/minima) or mean values of clinical indicators over one heart cycle.

Two technological trends make the present contribution particularly timely. On one hand, there is increasing importance attributed to the availability of large training datasets which is at the base of the current revolution in AI and deep learning [35]. This includes anonymous electronic health records (EHRs) for specific sub-populations affected by a common clinical condition. On the other hand, there is increasing availability of computational resources on the cloud which create a perfect infrastructure for distributed computing with lightweight models. For these reasons, we envision an increased adoption of numerical models as *regularizers* to determine physics-informed predictive distributions for missing data in EHRs, going beyond currently adopted, physics-agnostic multiple imputation methods [36]. This study aims to be a first step in this direction, and provides the following new contributions:

- We propose a systematic approach to train patient-specific circulatory models with clinical data uncertainty, and demonstrate the results obtainable on a modest cohort of 82 patients.
- Explore optimal parameter training as a possible approach to increase the feature space in order to facilitate classification of cardiovascular anomalies.

In Section 2.1, we discuss the differential formulations of a compartmental circulation model for human adults, including circuit elements, a generic heart model, systemic and pulmonary compartments. This is followed by an analysis of two datasets in Section 2.2, the first used for validation, while the second containing EHRs for 82 patients. Our numerical investigation is articulated through answers to the questions formulated in Section 2.3, using the numerical algorithms and tools briefly introduced in Section 2.4 and 2.5. Results are highlighted in Section 3 and related to physiological admissibility of the selected model, ability to capture dysfunction mechanisms, sensitivity and identifiability of input parameters, predictive performance for pulmonary pressures, non-pulmonary target ranking and viability of model-based PH classifiers. Conclusions are discussed in Section 4. Finally, for convenience, a list of acronyms is provided in Table 1.

## 2 Materials and Methods

### Compliance with Ethical Standards

The study was classified as research not involving human subjects and approved on June 13th, 2019 by the Office of Research Compliance and Institutional Review Board at the University of Notre Dame under IRB#19-05-5371. All procedures performed in studies involving human participants were in accordance with the ethical standards of the institutional and/or national research committee and with the 1964 Helsinki declaration and its later amendments or comparable ethical standards. For this retrospective study formal consent is not required.

### Supplementary Material

Supplementary material for this paper is available at https://github.com/desResLab/supplMatHarrod20

### 2.1 Modeling Approach

Blood circulation in adults can be simulated through a *lumped parameter model* (sometimes also referred to as *zero-dimensional* or *0D* model). The foundation beneath a lumped parameter model lies in the equations that govern the voltage and current in an electrical circuit. Utilizing the hydrodynamic analogy [37], these equations stem from conservation principles where the voltage is corresponded by the pressure and the current by the flow. In this study, we consider patient-specific 0D representations containing seven compartments: the four chambers in a bi-ventricular heart, a compliant aortic arch, and the pulmonary and systemic circulations. The pulmonary compartment is represented through an RC circuit as shown in Figure 1.

**Figure 1:**
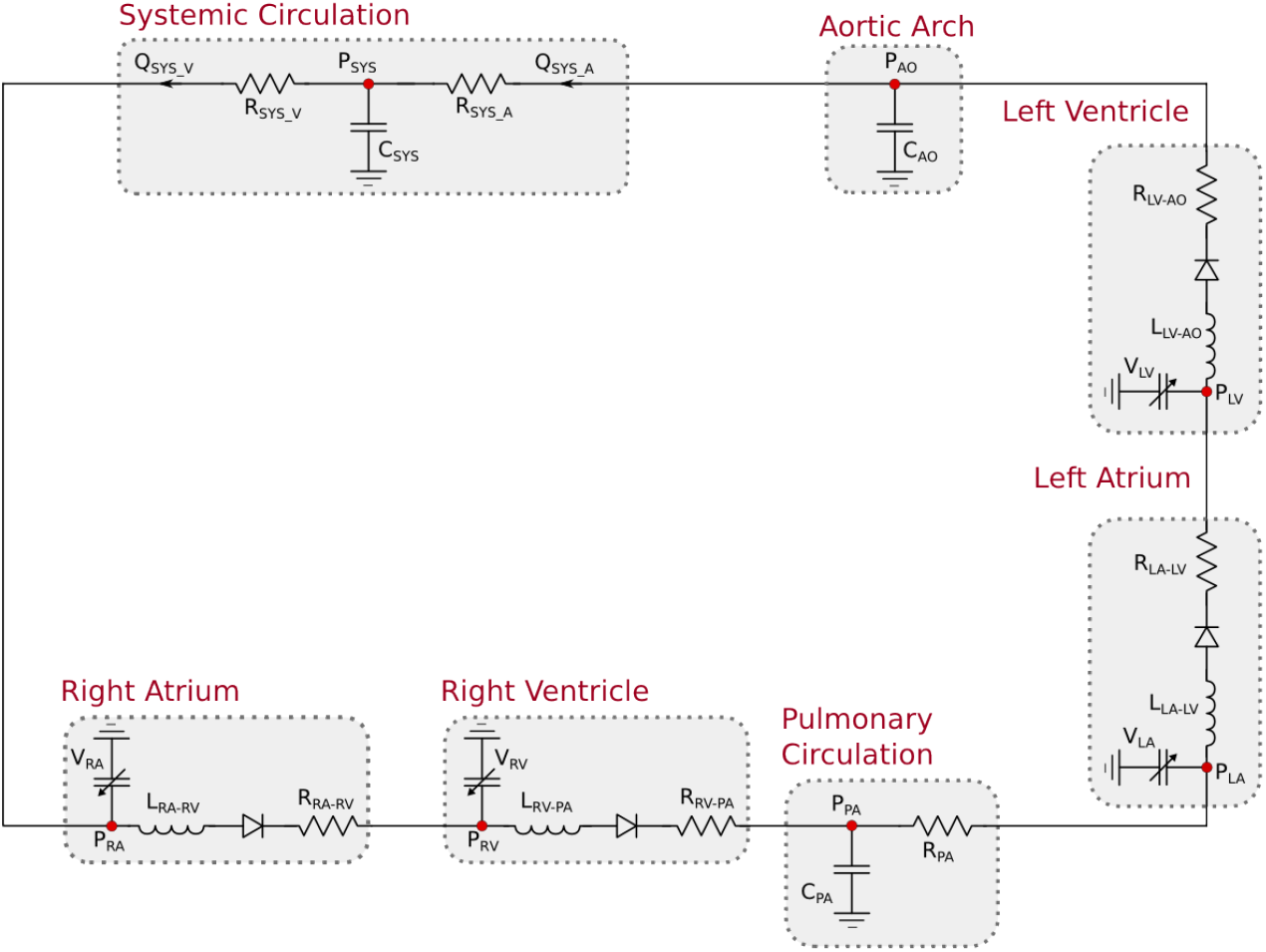
Lumped parameter hemodynamic model with RC pulmonary circuit.

#### 2.1.1 Generic Heart Model Compartment

The four heart chambers are represented by a series arrangement of a pressure-volume generator, an inductor, an ideal unidirectional valve and a resistor, following prior work. The atrial and ventricular pressure-volume generators are formulated through a combination of an activation function and active/passive pressure curves [10, 14]

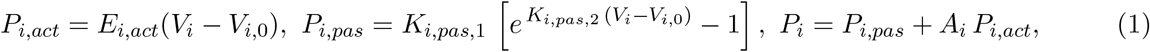

where the index *i* refers to either the right-left or atrial-ventricular chamber, *i* ∈ {*ra, rv, la, lv*}. The active pressure curve is assumed linear and characterized by an *active* elastance *E*_*i,act*_ and *unstressed chamber volume V*_*i*,0_. Additionally, passive volumes and pressures are related through an exponential relation, characterized by two elastance coefficients *K*_*i,pas*,1_ and *K*_*i,pas*,2_, respectively [10, 14]. The pressure-volume generator is completed by an *activation function A*_*i*_ of the form

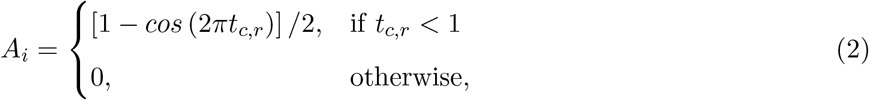

where *t*_*c,r*_ is the relative chamber activation time, ranging from 0 (beginning of systole) to 1 (end of systole). The chamber volume is also determined through the equations

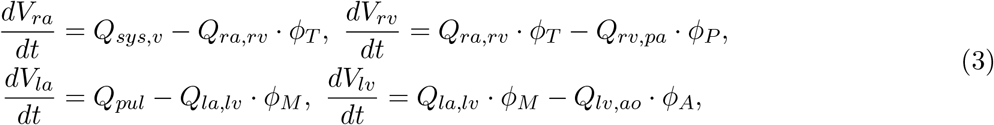

where *ϕ*_*i*_, *i* ∈ {*T, P, M, A*} are valve activation functions for the tricuspid (T), pulmonary (P), mitral (M) and aortic (A) valve respectively. These are equal to one for a negative pressure gradient through the valve and zero otherwise. Moreover, valves are modeled as *perfect*, without accounting for possible leakage or regurgitation. For models including valve prolapse and consequent regurgitation the reader is referred to, e.g., [38]. An inductance element located downstream with respect to the pressure-volume generator simulates the inertia of the blood in the chamber, according to the differential equation

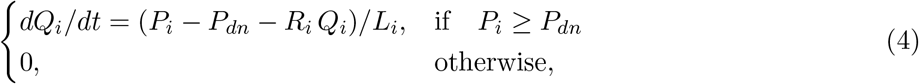

where *Q*_*i*_ is the volumetric blood flow going through the *i*-th chamber, *P*_*i*_ and *P*_*dn*_ the pressure in the *i*-th and downstream chamber, respectively, *R*_*i*_ the viscous resistance located between chamber *i* and chamber *dn*, and *L*_*i*_ an inductance parameter.

Additionally, the selected model is fully capable of representing the physiologic consequences of a stenotic valve, as an increase in the resistance of the associated compartment would accentuate the pressure drop across the valve and lead to an increase of the upstream pressure. Finally, we remark that systolic and diastolic functions are separately represented in this model, using three parameters for each chamber, i.e., *E*_*i,act*_, *K*_*i,pas*,1_ and *K*_*i,pas*,2_. Identification of these parameters from clinical health records would therefore be informative of systolic and diastolic chamber function.

Note how the RL parameters of each cardiac chamber are kept fixed in this study (see Table A1) or, in other words, we assume that valve stenosis has been excluded as a possible cause of pulmonary hypertension, for example through a non-invasive echocardiographic assessment.

#### 2.1.2 Aortic and Systemic Compartment

An aortic compartment consisting of an isolated capacitor is positioned downstream of the left ventricular outflow, modeled through an equation of the form

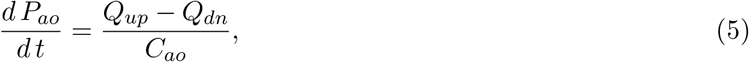

where *Q*_*up*_ and *Q*_*dn*_ are the volumetric flow rate from the left ventricle and abdominal aorta, respectively, *P*_*ao*_ is the aortic pressure and *C*_*ao*_ the aortic compliance. We compare the value of *P*_*ao*_ computed by this model with the clinically acquired brachial pressure.

An RCR circuit simulates the systemic circulation, with *C*_*sys*_ used to represent the overall systemic compliance, while two resistors simulate the viscous resistance in arteries and veins *R*_*sys,a*_ and *R*_*sys,v*_, respectively. The algebraic-differential equations for the systemic compartments are therefore:

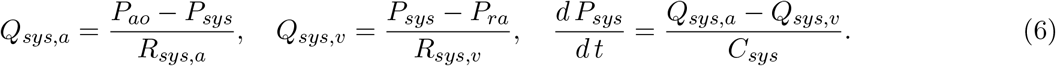

#### 2.1.3 Pulmonary Compartment

The pulmonary circulation is represented through a RC circuit with equations

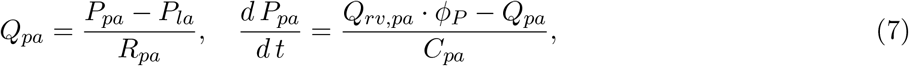

where the pulmonary, left atrial and right ventricular pressures are denoted by *P*_*pa*_, *P*_*la*_ and *P*_*rv*_, and pulmonary capacitance and resistance are *C*_*pa*_ and *R*_*pa*_, respectively. The pulmonary flow rate is denoted as *Q*_*pa*_, while *Q*_*rv,pa*_ denotes the flow across the pulmonary valve, having activation equal to

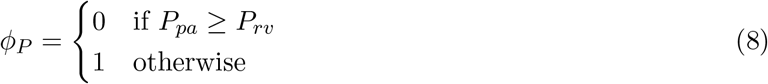

### 2.2 Available Data Sets

Two data sets are used throughout this study. Synthetic patient-agnostic clinical measurements representing increasing severity of diastolic left ventricular dysfunction are used initially, while anonymized patient-specific electronic health records (EHR) for a cohort of 82 patients are utilized in the second part of the study.

#### 2.2.1 Validation Data Set

The validation data set (Table 2) contains the mean and standard deviation of thirteen different clinical targets for three different heart failure groups: healthy patients, mild heart failure patients, and patients with severe heart failure. Normal physiologic clinical targets were determined from the literature [39], while targets associated with severe left ventricular diastolic dysfunction were assigned with the supervision of a clinician. The values for mild heart failure patients were obtained through a linear interpolation between severe dysfunction and healthy conditions. The selected targets for severe HF conditions are characterized by a normal SBP and DBP accompanied by a slight reduction in CO. These are designed to represent conditions where the patient is asyntomatic, consistent with the aim of the proposed approach, related to early disease detection. Situations characterized by decompensated heart failure are therefore excluded on purpose by the dataset in Table 2.

**Table 2:** Validation data set containing the mean and standard deviation of clinical targets for three levels of increasing diastolic heart failure severity.

| Qty | Description | Units | Severe HF | Moderate HF | Healthy | $\sigma$ |
| --- | --- | --- | --- | --- | --- | --- |
| HR | Heart Rate | bpm | 80 | 80 | 80 | 3 |
| RAP | Right Atrial Pressure | mmHg | 15 | 9 | 4 | 0.5 |
| sPAP | Systolic Pulmonary Artery Pressure | mmHg | 50 | 35 | 20 | 1 |
| dPAP | Diastolic Pulmonary Artery Pressure | mmHg | 25 | 19 | 12 | 1 |
| PCW | Pulmonary Capillary Wedge Pressure | mmHg | 25 | 17 | 9 | 1 |
| SBP | Systolic Blood Pressure | mmHg | 120 | 120 | 120 | 1.5 |
| DBP | Diastolic Blood Pressure | mmHg | 80 | 80 | 80 | 1.5 |
| SVR | Systemic Vascular Resistance | dynes·s·cm<sup>-5</sup> | 1800 | 1575 | 1350 | 50 |
| CO | Cardiac Output | L/min | 3.5 | 4.375 | 5.25 | 0.2 |
| sRV | Systolic Right Ventricular Pressure | mmHg | 50 | 35 | 20 | 1 |
| RVEDP | Right Ventricular End Diastolic Pressure | mmHg | 15 | 9 | 4 | 1 |
| sLV | Systolic Left Ventricular pressure | mmHg | 120 | 120 | 120 | 1.5 |
| LVEDP | Left Ventricular End Diastolic Pressure | mmHg | 25 | 16 | 6 | 2 |

#### 2.2.2 EHR Data Set

Completely anonymized patient-specific clinical measurements for 82 adult patients were provided in the context of a research project funded by Google through its ATAP initiative, focusing on *Modeling Noninvasive Measurements of Cardiovascular Dynamics*. There are 26 clinical data targets, which are listed below in Table 3. Missing data were present with the pattern highlighted in Figure 2, with patients having zero to nineteen of the clinical targets. Two of the 84 patients in the dataset did not have any of the relevant clinical targets and were therefore excluded from the study (see the two patients with zero available targets in Figure 2a). The remaining 82 patients had between one and nineteen clinical targets. While there is not a single patient that has all 26 measurements, at least three clinical targets (heart rate, diastolic blood pressure, and systolic blood pressure) are available for all but one patient. Finally, the standard deviations for each clinical target are also shown in Table 3, which were determined through a preliminary literature review [40, 41, 42].

**Table 3:**
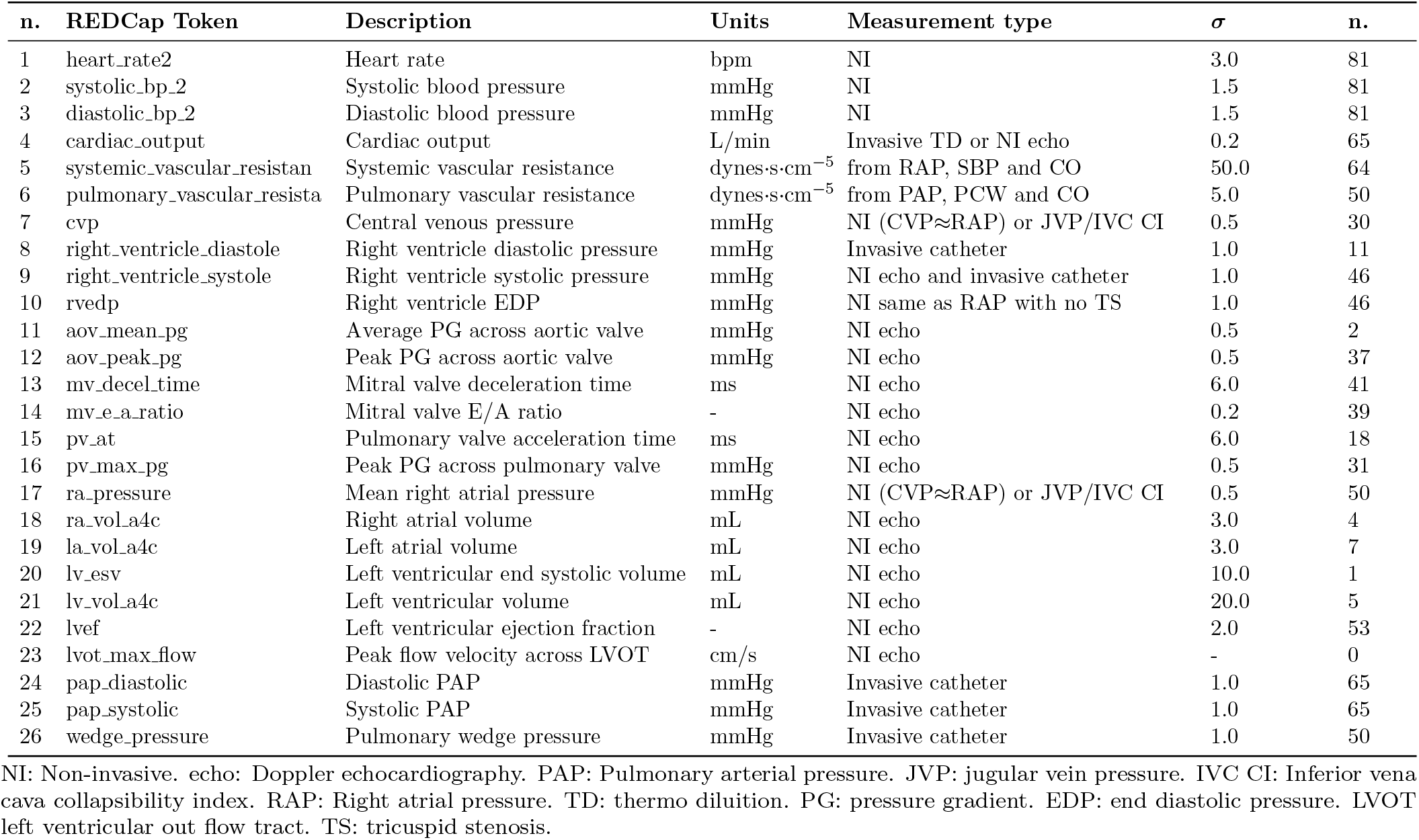
Patient-specific EHR data set containing 26 clinical measurements and associated units.

**Figure 2:**
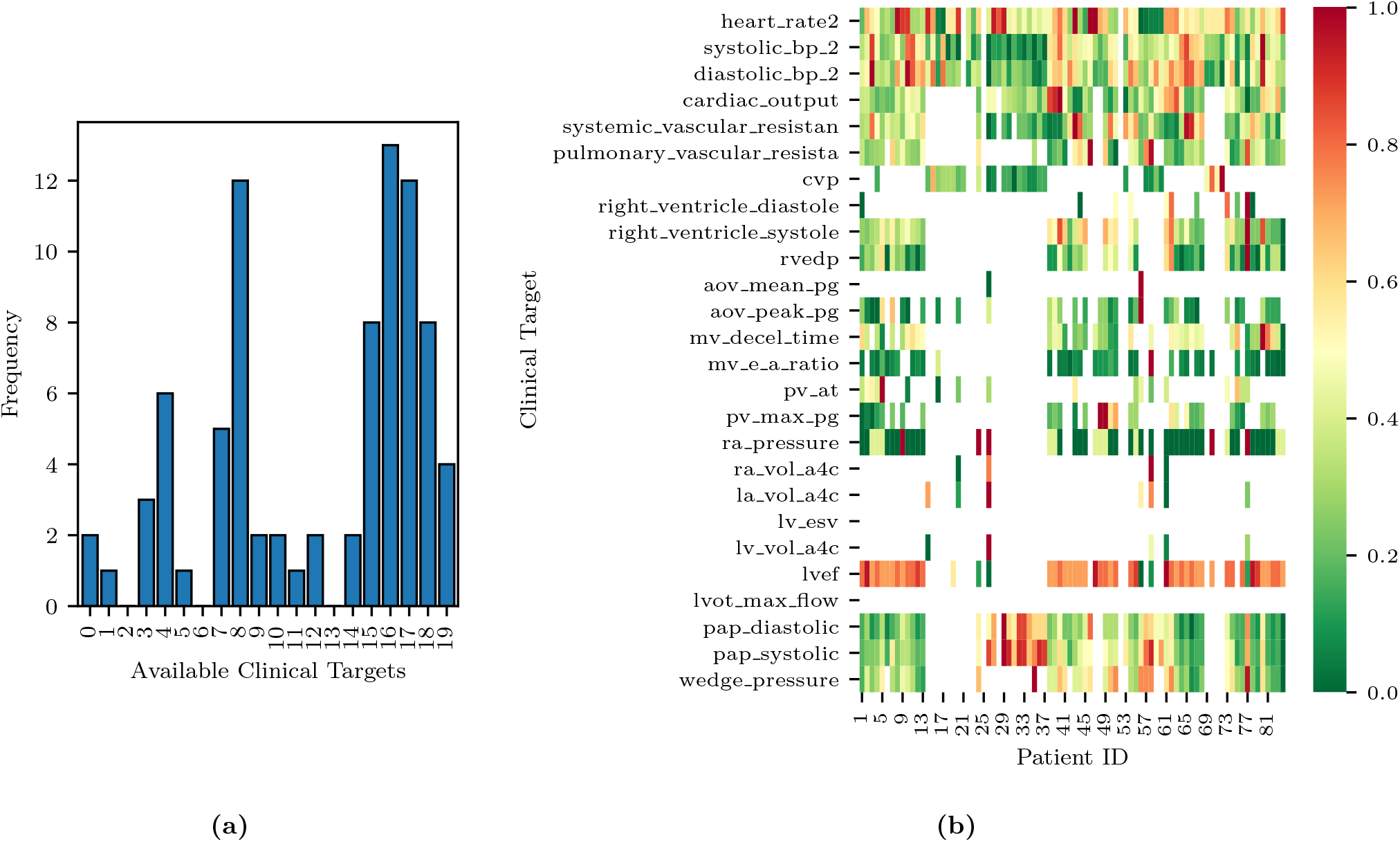
Histogram of data availability among the 82 patients (left) and missing data pattern (right). Note how each row of the heat map is normalized to a zero to one range, to highlight the relative magnitude of the clinical target.

A histogram of clinical data occurrences is illustrated in Figure 2a, and displays three main modes: the majority of patients have either four, eight, or seventeen available clinical measurements, likely due to the data aggregation produced by certain screening procedures. Additionally, a heat map of the EHRs is illustrated in Figure 2b, where each row of the heat map was normalized to a zero to one range, to highlight the relative magnitude of the clinical target.

Note how the size of the selected cohort (84 patients) is modest, but sufficient to investigate the effectiveness of Bayesian inference in the context of a simple physiologic model. Acquisition of larger data sets is possible but made non trivial by the need to automatically extract large volumes of clinical targets from text reports written following echocardiographic and catether lab examinations. In such cases, use of natural language processing tools is key to make larger EHR data sets available for research.

Finally, this data set focuses on cases of secondary PH, where a reversible increase of PVR follows an increase in left ventricular filling pressures. Therefore, this study does not consider primary PH and does not make any claim of differentiating primary from secondary PH.

### 2.3 Methodological Approach

This study is articulated through a number of logically consequential questions driving our numerical experiments. These questions are:

1. **Physiological admissibility of 0D representations under normal and heart failure conditions** - *Are model outputs able to reproduce sets of clinical targets ranging from healthy to pathological conditions?* In other words, is the identification problem *well-posed*, in the sense that model outputs are able to represent a wide spectrum of conditions from health to disease? We answer this question in Section 3.1.
2. **Ability to model distinct diastolic/systolic dysfunction mechanisms** - *Is the selected model formulation able to separately represent the systolic and diastolic functions of the heart muscle?* And does the alteration of these properties produce expected modifications in the physiology represented through model outputs? We answer this question in Section 3.2.
3. **Parameter sensitivity and identifiability** - Once a set of quantities whose prediction is of interest (e.g., pulmonary arterial pressure) has been identified, *do they show non-negligible sensitivity with respect to changes in the parameters associated with physiologic relevant mechanisms affecting these quantities*? Moreover, are these parameters *identifiable* so that it is possible to uniquely estimate their distribution from the available clinical data? Is this estimate robust (i.e., characterized by a limited uncertainty)? We answer this question in Section 3.3.
4. **Predictive ability of optimally trained models** - *Are models trained from clinical data other than the pulmonary pressures able to predict such pressures, and what is their accuracy* ? We answer this question in Section 3.4.
5. **Relative importance of non-pulmonary clinical targets** - *Which minimal set of clinical targets should be collected to guarantee accurate model predictions for systolic, diastolic pulmonary pressure and pulmonary venous wedge pressure*? In other words, we would like to *rank* the non-pulmonary targets, starting with those producing maximally accurate predictions on the pulmonary arterial pressure. This allows for identification of a minimal set of maximally informative clinical quantities for predicting specific model outputs. We answer this question in Section 3.5.
6. **Detecting pulmonary arterial hypertension from assimilated circulation models** - Group II pulmonary arterial hypertension is typically detected by a mean pulmonary pressure higher than 25 mmHg or a systolic pulmonary pressure higher than 35 mmHg [43]. *Instead of direct characterization based on clinical data, would it be possible to detect pulmonary arterial hypertension by classification from the parameters of a model trained with non-invasive measurements* ? Once assimilated, a model can be used to generate a large number of features, leading to a higher dimensional space with possibly improved separability [44]. We answer this question in Section 3.6.

### 2.4 Inference

Consider a set of *m* measured clinical targets represented through the random vector **d** ∈ ℝ^*m*^ with each component *d*_*i*_ ∼ *ρ*_*i*_(*d*_*i*_), *i* = 1, …, *m* and joint density **d** ∼ *ρ*(**d**) = *ρ*_1_(*d*_1_) *ρ*_2_(*d*_2_) … *ρ*_*m*_(*d*_*m*_). In other words, each quantity *d*_*i*_, *i* = 1, …, *m* measured in the clinic has marginal density *ρ*_*i*_(*d*_*i*_), *i* = 1, …, *m*, and we assume all these measurements to be independent, i.e., their joint probability factors. We design a physiologic 0D circuit model with *n* parameters **y** ∈ ℝ^*n*^, so its outputs match the observed targets or, in other words, we introduce a statistical model of the form

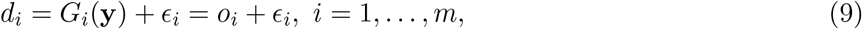

and assume each noise component 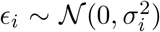 to follow a zero-mean Gaussian distribution. Note that the *i*-th realization from the parameter vector **y** is denoted by **y**^(*i*)^ and that the *i*-th model output is denoted by *o*_*i*_ = *G*_*i*_(**y**), while the vector **o** ∈ ℝ^*m*^ contains the complete set of model outputs.

Each model is *trained* using two different approaches, i.e., by determining a maximum a posteriori estimate of the parameters **y** using repeated Nelder-Mead optimization [45], and by solving an inverse problem through adaptive Markov chain Monte Carlo sampling, specifically, through the differential evolution adaptive Metropolis algorithm [46, 47] and assessing convergence through the Gelman-Rubin diagnostic [48]. In both cases, the posterior distribution *P* (**y**|**d**) is obtained by combining a uniform prior *P* (**y**) (see the admissible parameter ranges in Table A1) with a Gaussian likelihood

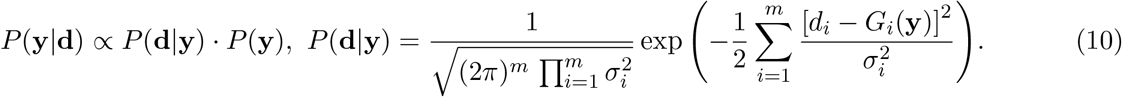

### 2.5 Computational tools

The tulip (Tools for Uncertainty quantification, Lumped modeling and Identification of Parameters) software framework was developed to answer the above research questions. Tulip is a OOP C++ code designed to simplify the task of estimating parameters of lumped models for human circulation and contains abstractions for computational models, operations performed on these models (e.g., optimization, Bayesian estimation, local and global sensitivity analysis, etc.) and data sources used to store the available clinical targets. For an overview of the procedures for statistical data assimilation used in this study, the interested reader is referred to [49, 50].

## 3 Results

### 3.1 Physiological admissibility

The circulation model discussed in Section 2.1 was first trained on the validation dataset in Section 2.2.1. We then generated a collection of model outputs using a subset **y**^(*i*)^, *i* = 1, …, 5000 parameter realizations from the converged MCMC samples, and compared the resulting distributions (after post-processing with Gaussian kernel density estimation) with the distributions assumed for the targets **d**. Specifically, we assumed that each clinical target follows a normal distribution with the mean and standard deviations listed in Table 2. The Kullback-Leibler (KL) divergence [51] was used to determine the agreement between the model-based predictions and measurements. The KL divergence measures how much information is lost when one uses the predicted, instead of the assumed, target distribution and a small KL divergence suggests physiological admissibility. As shown in Figure 3, the KL divergence is negligible for all targets, and the relative percent difference between mean target value and MAP model outputs is also generally small (see Table 4).

**Table 4:** Validation data set. Percent differences between model outputs for MAP parameter estimates and clinical targets for various degrees of heart failure severity.

|  | Healthy |  |  | Mild |  |  | Severe |  |  |
| --- | --- | --- | --- | --- | --- | --- | --- | --- | --- |
|  | Target | Computed | Error [%] | Target | Computed | Error [%] | Target | Computed | Error [%] |
| HR | 80 | 80.8 | 0.94 | 80 | 80.6 | 0.81 | 80 | 81.3 | 1.63 |
| RAP | 4 | 4.1 | 1.70 | 9 | 9.0 | 0.21 | 15 | 15.0 | 0.19 |
| sPAP | 20 | 18.7 | 6.59 | 35 | 33.0 | 5.70 | 50 | 49.3 | 1.49 |
| dPAP | 12 | 11.9 | 0.84 | 19 | 18.6 | 2.34 | 25 | 24.4 | 2.22 |
| PWP | 9 | 9.1 | 1.65 | 17 | 16.4 | 3.80 | 25 | 25.1 | 0.34 |
| SBP | 120 | 118.3 | 1.43 | 120 | 116.6 | 2.84 | 120 | 117.4 | 2.15 |
| DBP | 80 | 79.5 | 0.61 | 80 | 78.4 | 1.96 | 80 | 78.9 | 1.37 |
| SVR | 1350 | 1397.4 | 3.51 | 1575 | 1594.8 | 1.26 | 1800 | 1819.7 | 1.09 |
| CO | 5.25 | 5.4 | 2.62 | 4.375 | 4.4 | 0.28 | 3.5 | 3.6 | 2.51 |
| sRV | 20 | 21.4 | 6.79 | 35 | 35.4 | 1.15 | 50 | 50.0 | 0.02 |
| dRV | 4 | 4.2 | 4.96 | 9 | 8.5 | 5.46 | 15 | 15.2 | 1.27 |
| sLV | 120 | 121.4 | 1.18 | 120 | 118.9 | 0.94 | 120 | 119.9 | 0.07 |
| dLV | 6 | 4.9 | 19.04 | 16 | 15.1 | 5.38 | 25 | 23.1 | 7.50 |

**Figure 3:**
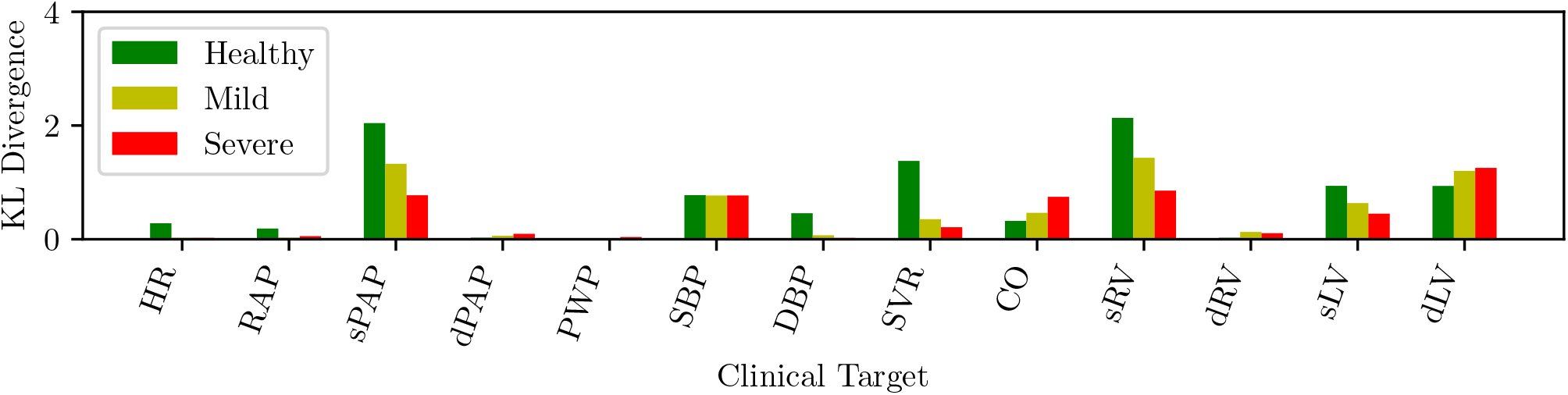
KL divergence between predicted and assumed clinical targets for varying heart failure severity.

### 3.2 Inverse assessment of dysfunction mechanisms

We now focus on the ability of our model to distinguish between diastolic and systolic dysfunction mechanisms when trained based on conditions reported in Table 2. Specifically, we aim to determine whether the selected model parameterization is able to separately represent the relaxation or contraction (i.e. diastolic or systolic) function of the heart muscle. Recall that diastolic function, and therefore left ventricular stiffness, during relaxation relate to the linear and exponential passive curve parameters *K*_*i,pas*,1_ and *K*_*i,pas*,2_ in equation (1). These two parameters directly relate to the condition that we aim to assess.

Figure 4 shows the mean diastolic and systolic chamber function parameters and associated 10%-90% confidence intervals, grouped by anatomical relevance (i.e., left ventricle, right ventricle) and plotted for healthy patients and patients with mild and severe heart failure, respectively. Comparison between heart failure conditions allows one to assess how model parameters change as a result of disease progression. This change is shown in Figure 4, where many of the parameters that relate to pulmonary hypertension do in fact change as patients progress from healthy to mild, and mild to severe diastolic dysfunction. The results show how the passive left ventricular curve slope *K*_*lv,pas*,1_ increases when going from healthy to mild and mild to severe, while the pulmonary resistance increases, as expected. Additionally, the left ventricular active elastance parameter *E*_*max,lv*_ remains nearly constant; the model correctly predicts no reduction in systolic function. This confirms that the physiological model used for this study can in fact distinguish between systolic/diastolic dysfunction mechanisms. Finally, the significant variability in the unstressed ventricular volumes is explained by the presence of multiple local peaks in the posterior distribution, i.e., reasonable PV loops compatible with the clinical targets produced by wildly different unstressed volumes.

**Figure 4:**
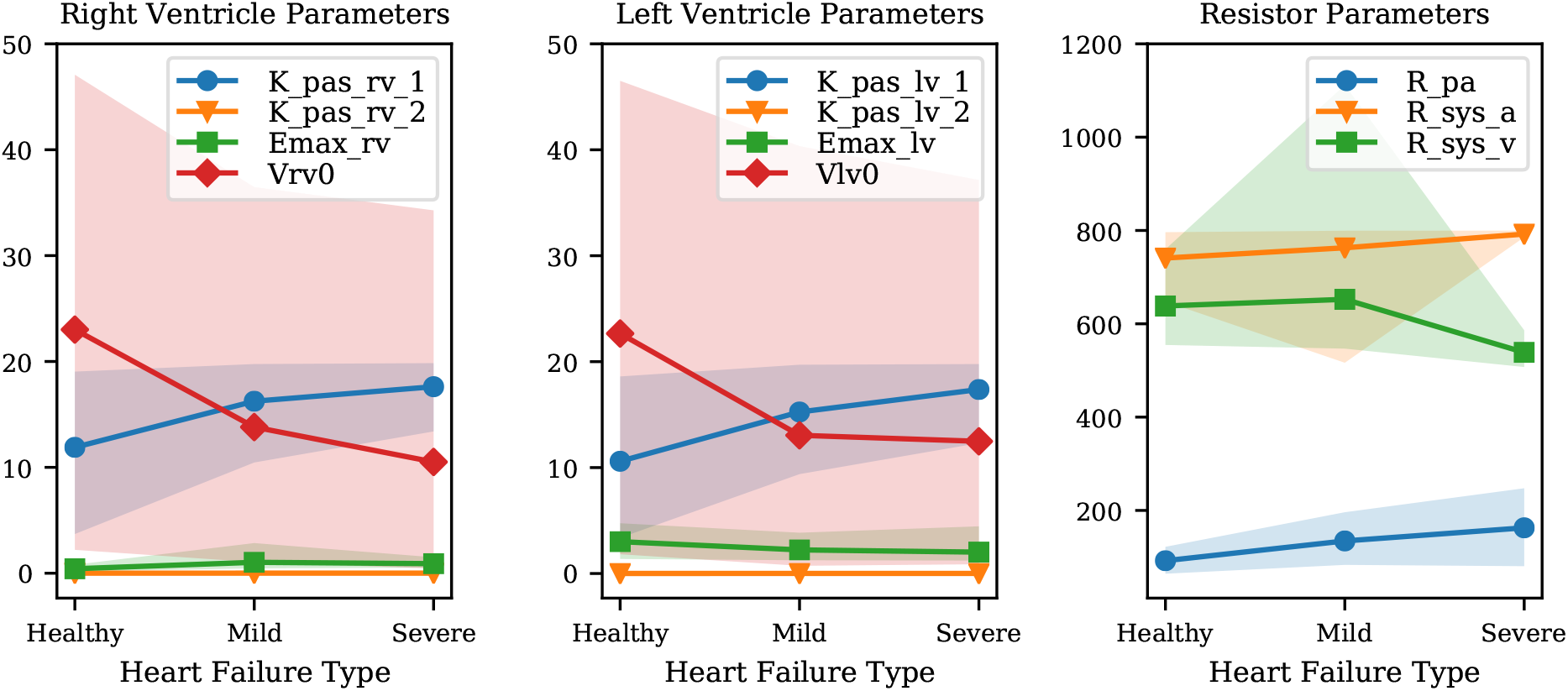
Variation of mean parameter values and associated confidence intervals under increasing heart failure severity. Right and left ventricular model parameters shown in the Figure are the ventricular passive curve slope *K*_*pas,i*,1_, exponent factor *K*_*pas,i*,2_, active curve slope *E*_*max,i*_ and unstressed ventricular volume *V i*, 0 (where *i* ∈ {*rv, lv*}). It also shows the following resistance parameters: *R*_*pa*_ (pulmonary resistance), *R*_*sys,a*_ (arterial systemic resistance), *R*_*sys,v*_ (venous systemic resistance).

### 3.3 Sensitivity and identifiability of circulation model parameters

Results from the above sections confirm the *well-posedness* of the selected model formulation for the full spectrum of diastolic dysfunction, from mild to severe. We now focus on determining the most relevant model parameters that significantly alters the main quantities of interest, particularly the systolic, diastolic and pulmonary wedge pressures. In addition, we study both the structural and practical identifiability in an effort to determine unimportant parameters and their non-identifiable combinations.

#### 3.3.1 Average local sensitivities

Nondimensional local sentitivities are computed for all outputs as

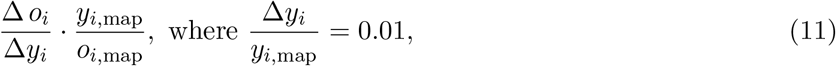

so that we consider the relative change in model outputs that correspond to a 1% variation in each parameter. The maximum a posteriori parameter vector **y**_map_ and the corresponding model outputs **o**_map_ = **G**(**y**_map_) are computed from MCMC for each of the 82 patients in the cohort, and used to compute the sensitivities in equation (11). The resulting sensitivities are then averaged across all patients.

Figure 5a illustrates the average sensitivities obtained by training our model with the complete list of clinical targets, including systolic, diastolic and venous wedge pulmonary pressures and pulmonary vascular resistance. We note that large sensitivities are apparent for the heart rate across all outputs while at the same time, accurate measurements of heart rate are easy to obtain non-invasively. Additionally, to check how the sensitivities were affected by the availability of pulmonary pressure targets (i.e., the very same quantities we would like to predict), we re-computed sensitivities using parameter estimates **y**_map_ obtained by excluding the pulmonary pressure targets during training. The average sensitivities appear to be minimally affected by the selective inclusion of pulmonary pressure targets.

**Figure 5:**
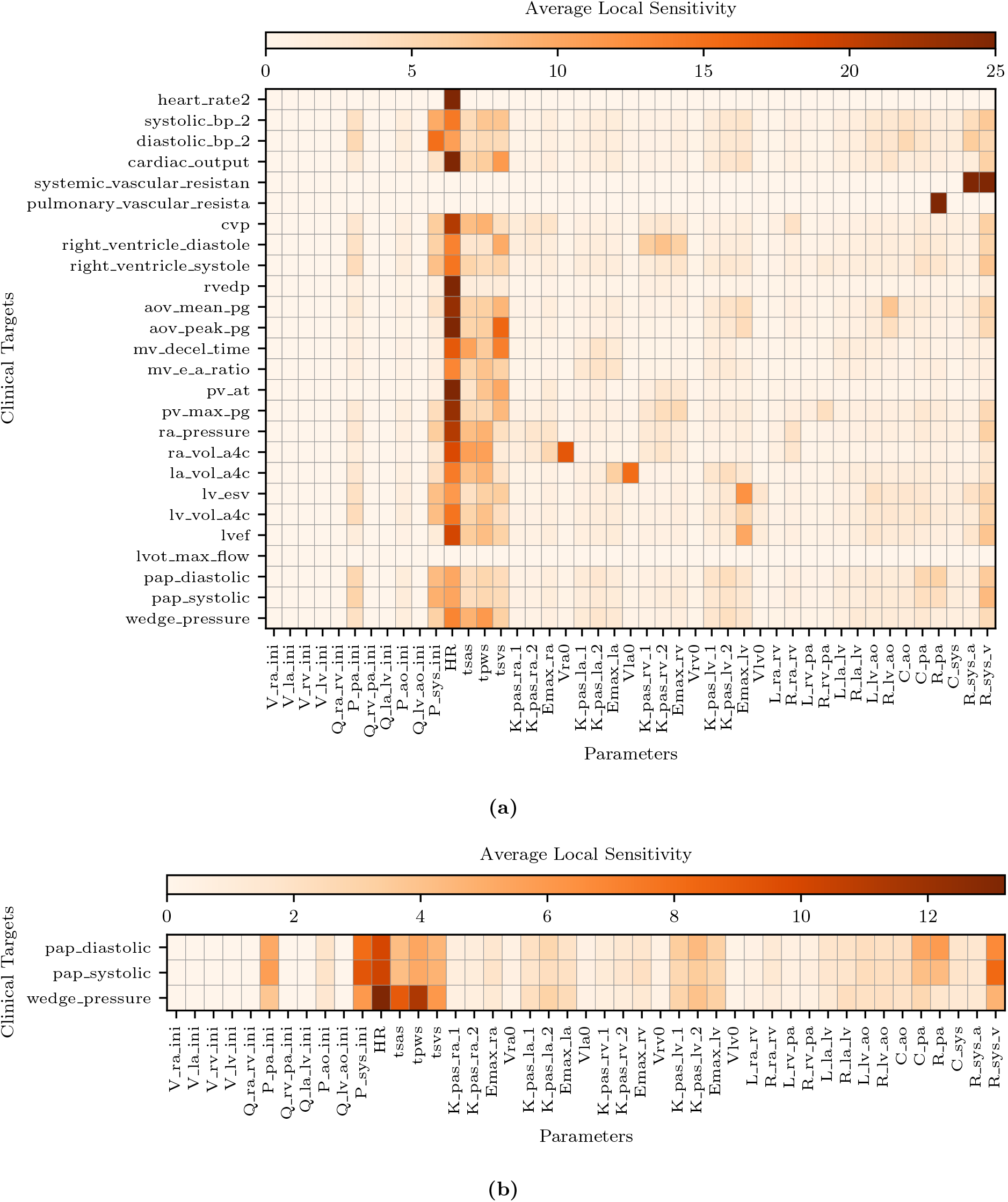
Average local sensitivities using a perturbation factor of 1%. (a) Average local sensitivity table for all parameters and model outputs. (b) Average local sensitivities table for all parameters and pulmonary outputs only. The values in each row are scaled to make their sum equal to 100.0 and a two cutoffs are used equal to 25 and 12.5 for (a) and (b), respectively.

**Figure 6:**
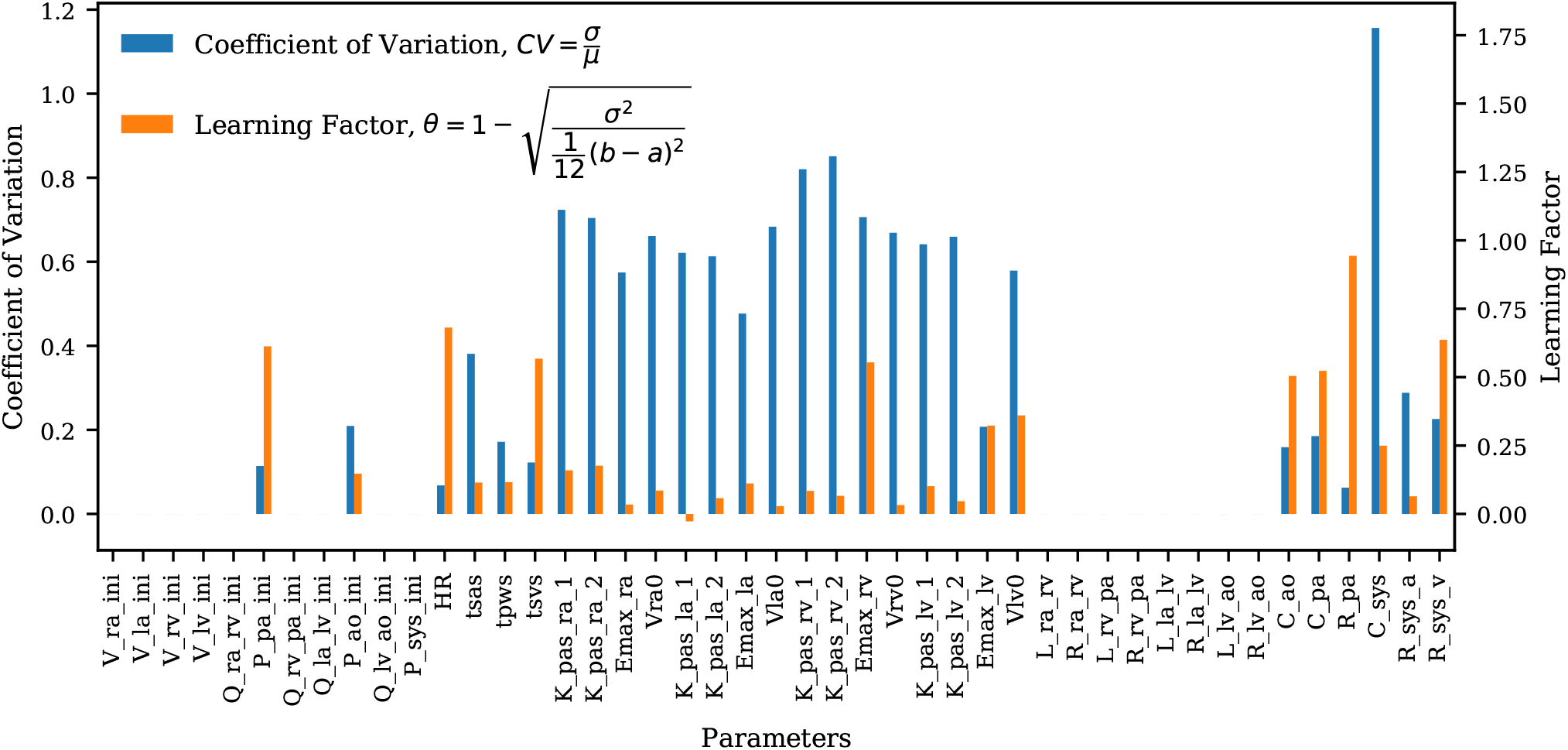
Bar plot of the coefficient of variation (*CV*) alongside the learning factor, (*θ*) for each parameter. *a* and *b* represent the minimum and maximum bound of each parameter uniform marginal prior.

Figure 5b suggests the following important parameters. Changes in the heart rate or *t*_*svs*_ directly affect the amount of blood flow ejected by the ventricles, altering the mean pulmonary pressures under a constant PVR and initial value (i.e., *P*_*pa,ini*_). As already discussed in the above sections, the left ventricular diastolic pressure/volume ratio *K*_*lv,pas*,1_ and the associated exponential factor *K*_*lv,pas*,2_ govern the diastolic properties of the left ventricle, while the *E*_*max,lv*_ is instead responsible for the systolic function. The left atrioventricular and aortic valve resistance *R*_*la,lv*_ and *R*_*lv,ao*_, respectively, govern the pressure drop from the left atrium to the left ventricle and from the left ventricle to the aorta. These two parameters therefore affect the left atrial and ventricular pressures and, in turn, the upstream pulmonary pressures. Mitral or aortic valve stenosis are typical examples of this mechanism (see, e.g., [52]). The pulmonary resistance and capacitance parameters *R*_*pa*_ and *C*_*pa*_ clearly affect the mean pulmonary pressures and their range, while changes in systemic vascular resistance *R*_*sys,a*_ or *R*_*sys,v*_ affect the left ventricular afterload and, in turn the pulmonary pressures.

#### 3.3.2 Structural identifiability

Structural identifiability analysis is performed to gain understanding of our ability to recover a given set of model parameters, through the solution of an inverse problem from *idealized noiseless* clinical targets that belong to the model range. This is in contrast with the analysis in the next section of the *practical* identifiability where real clinical data are used instead. We solve the model for the default parameter combination in Table A1 and regard the outputs as data, from which a MAP estimate of the default parameter values is re-computed by MCMC followed by NM optimization. We would like to point out how the parameters in Table A1 correspond to a *healthy* patient and therefore no attempt is made in this section to represent hypertensive physiological conditions. Additionally, histograms are generated using 5000 samples from the MCMC parameter traces and compared to the default (true) parameter set. The results for right, left ventricular and resistor parameters can be seen in Figures 7, 8, and 9, respectively. The true parameters are always found within the parameter distributions from MCMC and often correspond to the mode of the histogram. Some deviations may be observed in Figure 9, with reference to the arterial and venous systemic resistance, respectively. In such a case, since SVR is the sum of such resistances, the increase in the first is compensated by a reduction in the second.

**Figure 7:**
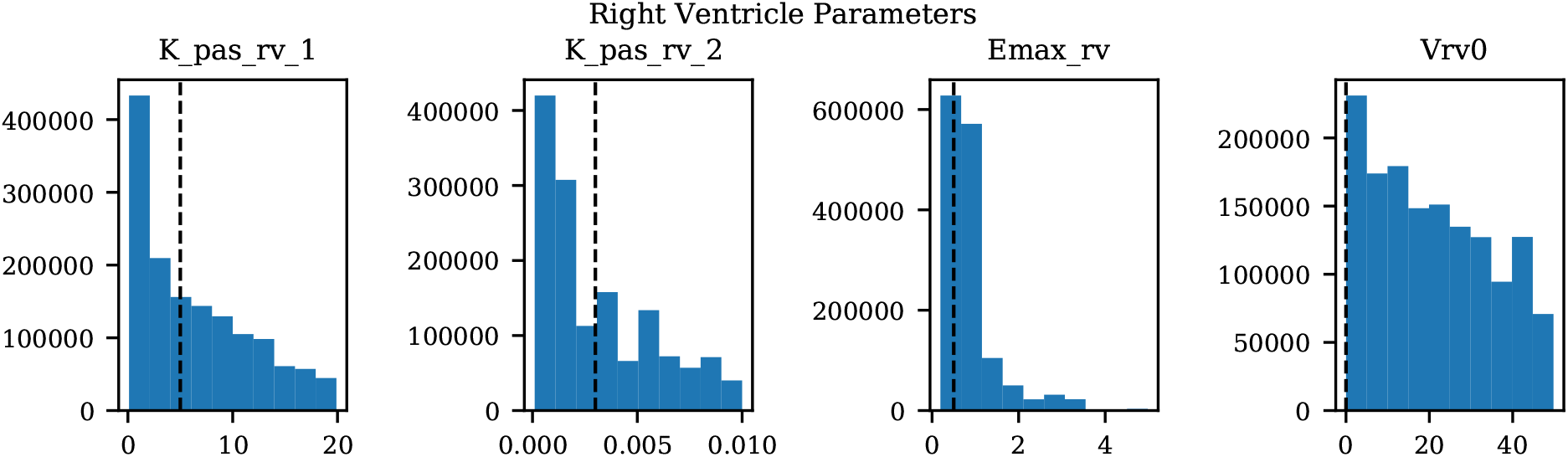
Histograms of right ventricular model parameters from MCMC and default parameter values used for assessing structural identifiability.

**Figure 8:**
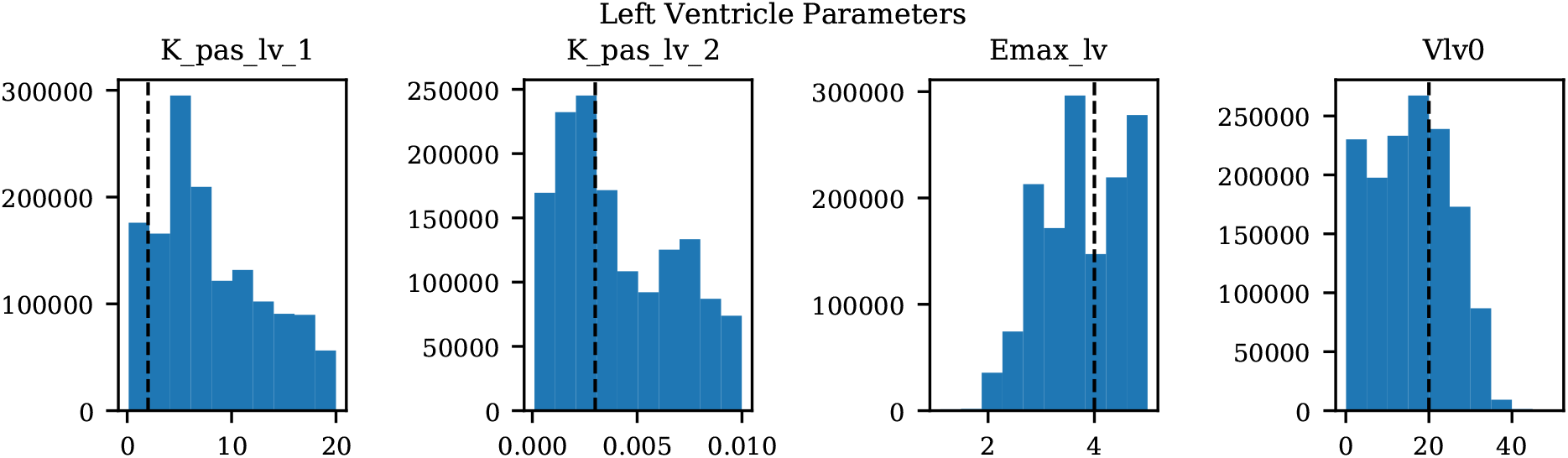
Histograms of left ventricular model parameters from MCMC and default parameter values used for assessing structural identifiability.

**Figure 9:**
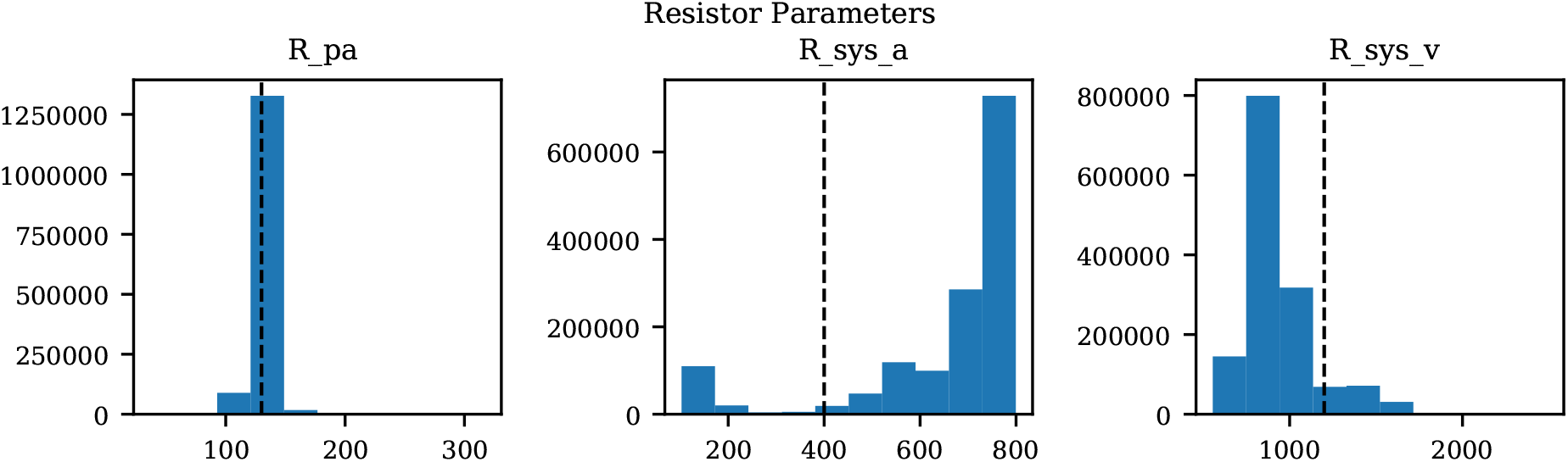
Histograms of pulmonary and systemic resistances from MCMC and default parameter values used for assessing structural identifiability.

The coefficients of variation for the model parameters marginal posteriors and associated *learning factors* are shown in Figure 6. While parameters with higher coefficient of variation have a greater spread relative to their mean, the learning factor quantifies how much the marginal variance is reduced by conditioning the model output to the available observations or in other words how much the marginal variance is reduced from the prior to the posterior [49]. Figure 6 shows that the parameters with the largest coefficient of variation are also the parameters with the smallest learning factor, as expected. Heart timing parameters, systemic and pulmonary resistance and compliance, aortic compliance and active ventricular parameters are generally well learned.

The analysis is completed by a comparison between the true and optimal physiology, calculated using 5000 parameter combinations from the MCMC traces, and analyzed over a single heart cycle. The results for the aortic, pulmonary, and left ventricular pressures and flows, as well as the left and right ventricular pressure-volume loop are reported in Figure 10. Pressures, flows and volumes agree well with those generated from the default parameter set.

**Figure 10:**
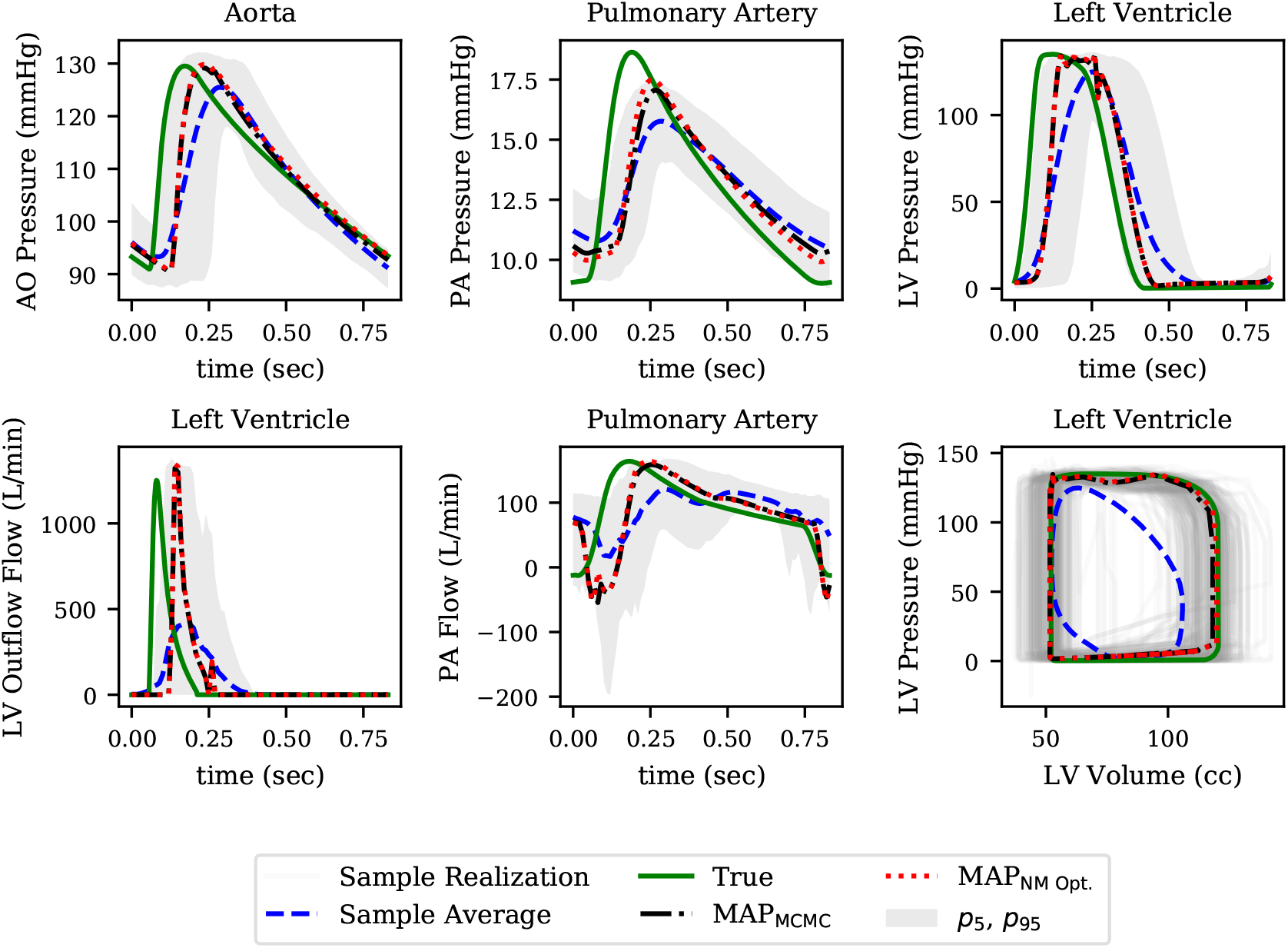
Pressure time histories over one heart cycle for aortic, pulmonary and left ventricular pressure (row 1). Flow time histories over one heart cycle for left ventricular outflow and pulmonary flow as well as left ventricular pressure-volume loop. (row 2).

#### 3.3.3 Practical identifiability

We also perform local identifiability analysis through the Fisher information matrix rank [53] to determine the presence of non-identifiable parameter combinations and unimportant parameters. To do so, we compute the matrix *∂* **G**(**y**_map_)*/∂* **y** of local derivatives for our output quantities **o**_map_ = **G**(**y**_map_) with respect to the parameters **y**. The Fisher Information matrix ℐ(**y**_map_) can be computed as

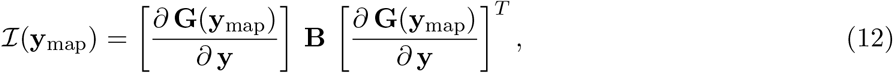

where the *precision* matrix **B** contains the inverse target variances, i.e., 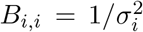 and *B*_*i,j*_ = 0 for *i* ≠ *j*. Rank deficiency in ℐ(**y**_map_) reveals the presence of non identifiable parameter combinations as illustrated in Figure 11a by plotting the Fisher information matrix (FIM) eigenvalues ordered by magnitude for all patients. Small eigenvalues are observed across all analyzed patients, confirming a lack of local identifiability.

**Figure 11:**
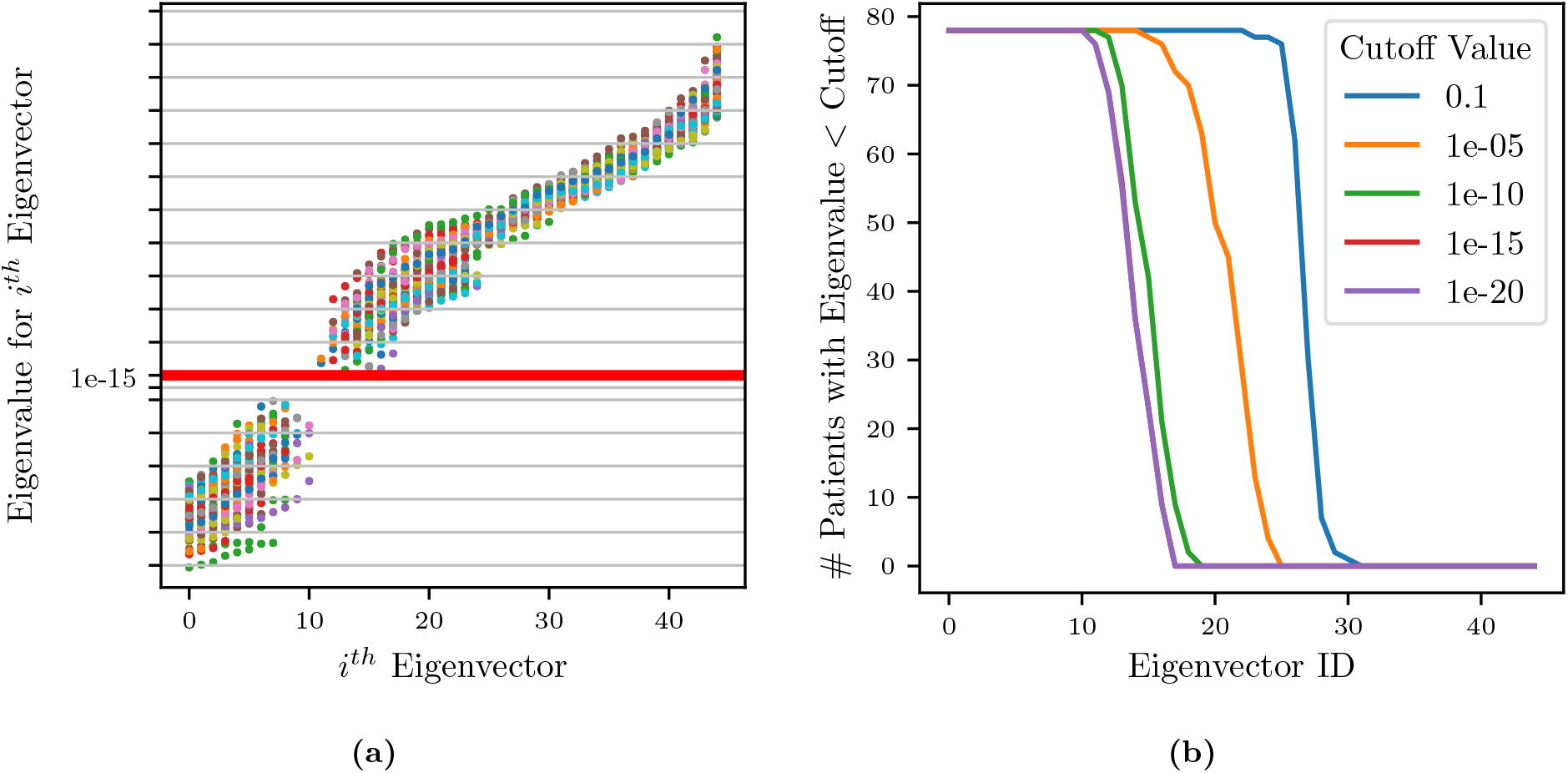
(a) Scatter plot of eigenvalues vs. eigenvectors for all patients. Red horizontal line represents selected cut-off value. FIM eigenvalues at identified parameters plotted in increasing order. (b) Plot of number of patients selected (eigenvalues less than cut-off) for each eigenvector.

Eigenvectors for all patients whose corresponding eigenvalues were less than a selected cut-off were superimposed on the same radar plot, to search for situations characterized by dominant components, representative of unimportant parameters. This process is visualized in Figure 11a, illustrating that all eigenvalues colored by patient and Figure 11b, showing the effect of changing the cut off value on the number of selected patients. Specifically, it shows that no significant changes result by adopting cut-offs in the range [1 × 10^*−*12^, 1 × 10^*−*16^].

Figure 12 shows an example of two of the 17 radar plots generated for this study. The plot on the left shows an example of unimportant parameter with dominant eigenvalue associated with the parameter *V*_*la,ini*_, i.e., the initial left atrial volume. Instead, the radar plot on the right does not show any clear pattern involving parameter combinations that significantly change across patients. This local identifiability analysis confirms how initial conditions for pressures, flows and volumes (except *P*_*pa,ini*_, *P*_*ao,ini*_ and *P*_*sys,ini*_ as per the results of the previous sensitivity analysis) are generally unimportant.

**Figure 12:**
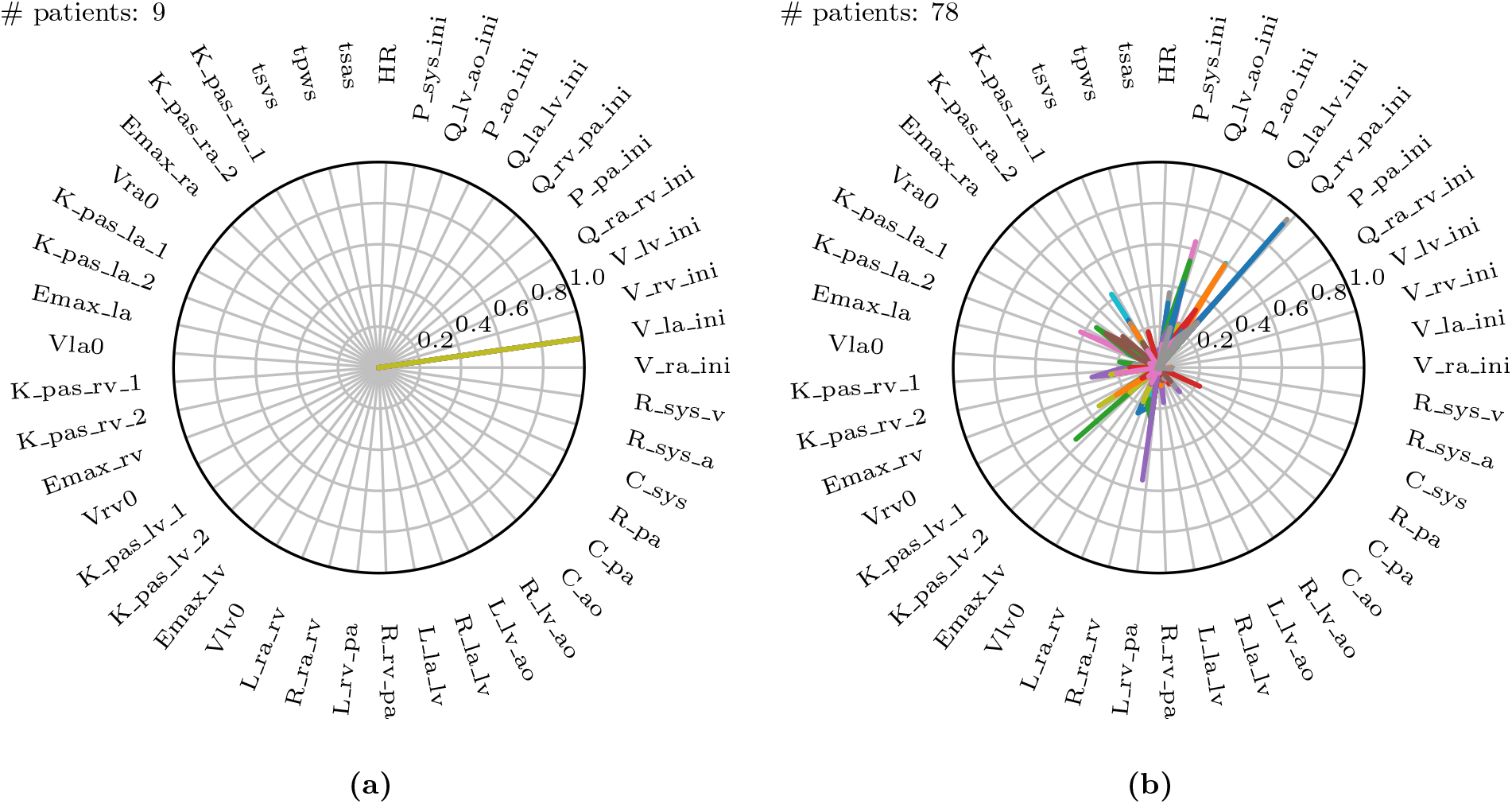
Selection of two of the 17 total radar plots of all parameters whose eigenvalues are less than the selected cut-off. (a) Example of unimportant initial condition, i.e. whose perturbation has no effect on the model results. (b) Example of non-identifiable parameter combinations where no dominant parameter can be identified and where the combination significantly changes across patients.

### 3.4 Prediction of pulmonary pressures

We now focus on the problem of using the physiological consistency of our compartmental model to *predict* pulmonary pressures from other possibly non-invasive clinical targets. To do so, we have trained our models without including the pulmonary pressures (i.e., systolic, diastolic, wedge pressures and pulmonary vascular resistance) as targets, and propagated forward the estimated parameters to quantify the marginal distributions for the predicted pulmonary pressures. We then evaluated the error between predicted and true pulmonary pressures together with their variability. To do so, we first show four Blant-Altman plots for sPAP, dPAP, mPAP and PVR, respectively. Model training including the pulmonary pressures and PVR generates very limited bias. Predictions generated by pulmonary pressure-blind training does not seem to generate proportional bias except for the predicted PVR.

In Figure 13c we also plot the average absolute pressure error across 31 patients (those for which the pulmonary pressure were available and characterized by having more than 6 REDCap entries) versus the minimum number of prescribed clinical targets, with the associated uncertainty. Finally, as illustrated in the closeup shown in Figure 13d which focuses on patients with at least 15 REDCap entries, the average errors on the predicted pressures is around 8 mmHg for systolic PAP, and 6 mmHg for Diastolic PAP and PCW.

**Figure 13:**
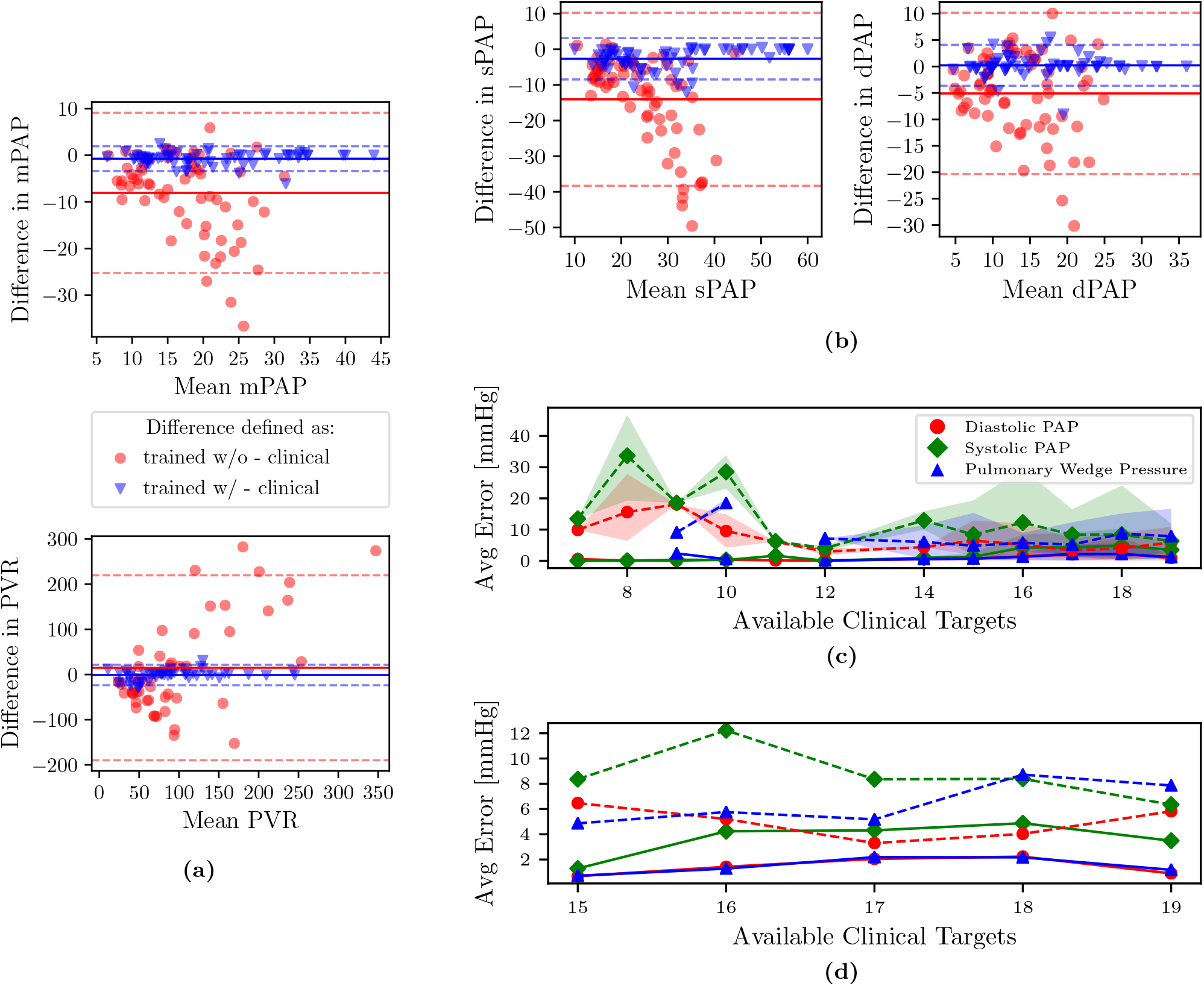
(a,b) Bland-Altman plots of sPAP, dPAP, mPAP and PVR for clinical targets, and as predicted from models trained with and without pulmonary targets, respectively. Solid horizontal lines represent the mean of all differences, while dashed lines are drawn at 1.96 times the standard deviation. (c) Predictive performance for pulmonary pressure. Absence of PAP targets in average prediction errors is represented using dashed lines. The shaded region represents the area bounded by the 5th and 95th percentile from 5000 random subsamples from MCMC. (d) Zoom on average errors which correspond to patients with more than 15 available clinical targets.

### 3.5 Relative importance of non-pulmonary targets

In this section, we investigate which clinical targets are the most important to include during training, in order to minimize errors in pulmonary pressure predictions. In other words, we rank clinical targets starting from those having a more beneficial impact on the accuracy in predicting pulmonary hypertension. We achieve this goal through a sequence of optimization steps. We start by performing training using optimization with a single target at a time. The target found to minimize the average *combined prediction error* for pulmonary pressures is ranked first and permanently added to the list of targets to be included in all successive optimization steps. The percent error was computed for each clinical pulmonary target that was available for each patient. Percentage error expresses the percentage difference between the model output and the clinical target, while a *cumulative* percentage error was calculated by taking the sum of the percentage errors for each pulmonary target that was clinically available for each patient. To avoid bias towards patients with fewer available pulmonary targets, the cumulative percentage error is transformed to an *average* combined prediction error through division by the number of available pulmonary targets. Let *p* ∈ {1, 2, 3, 4} be the number of pulmonary targets that is available in the data of a certain patient. The percent error of clinical target *s*_*i*,target_, *i* = 1, …, *p* is computed as *e*_*i*_ = 100 · |*s*_*i*,target_ − *s*_*i*,output_|*/s*_*i*,target_, *i* = 1, …, *p*, while the average combined prediction error is 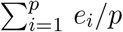.

All remaining targets are iteratively tested and those producing the minimum average combined PAP errors are progressively ranked, until all targets have been considered. The process above is repeated for each patient. Figures 14 and 15 show the resulting average ranking and associated occurrences, the number of patients where a specific target was collected.

**Figure 14:**
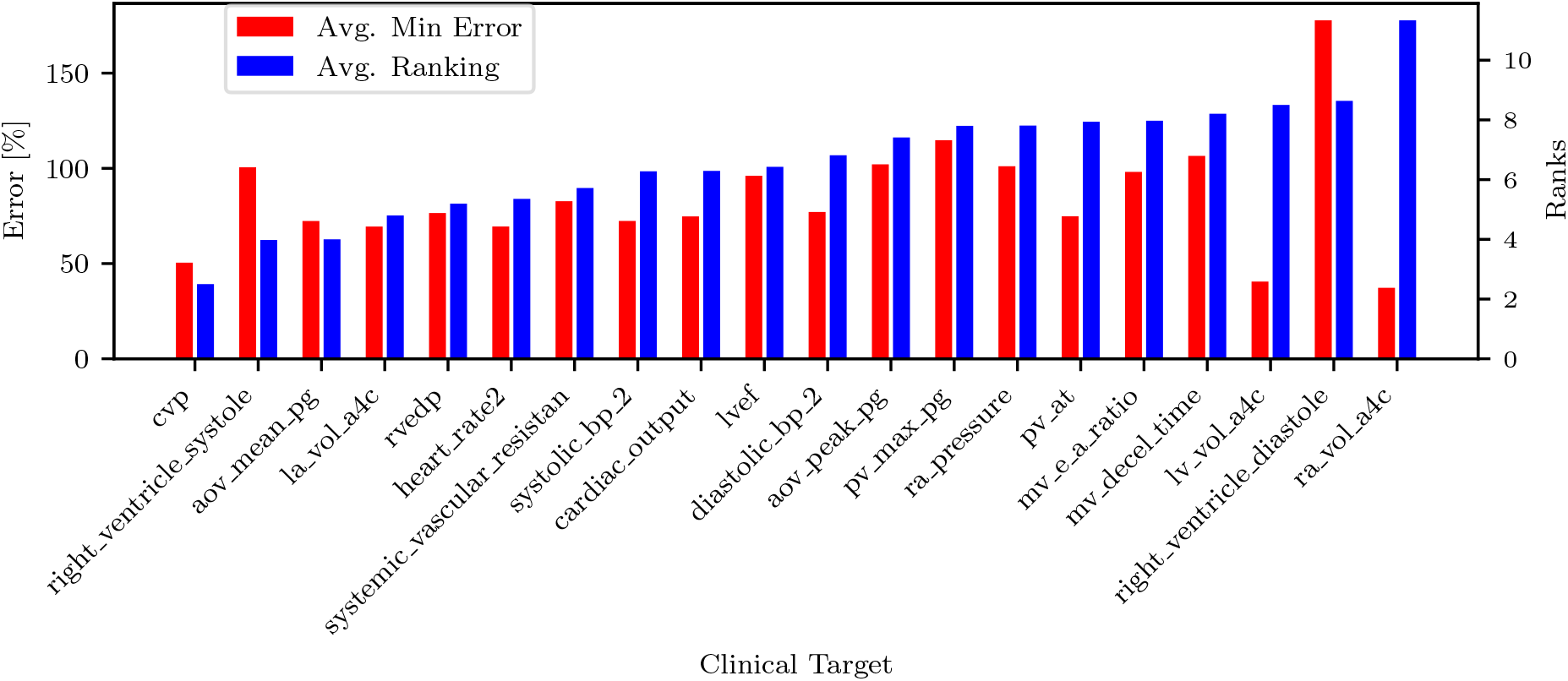
Target ranking and minimum error.

**Figure 15:**
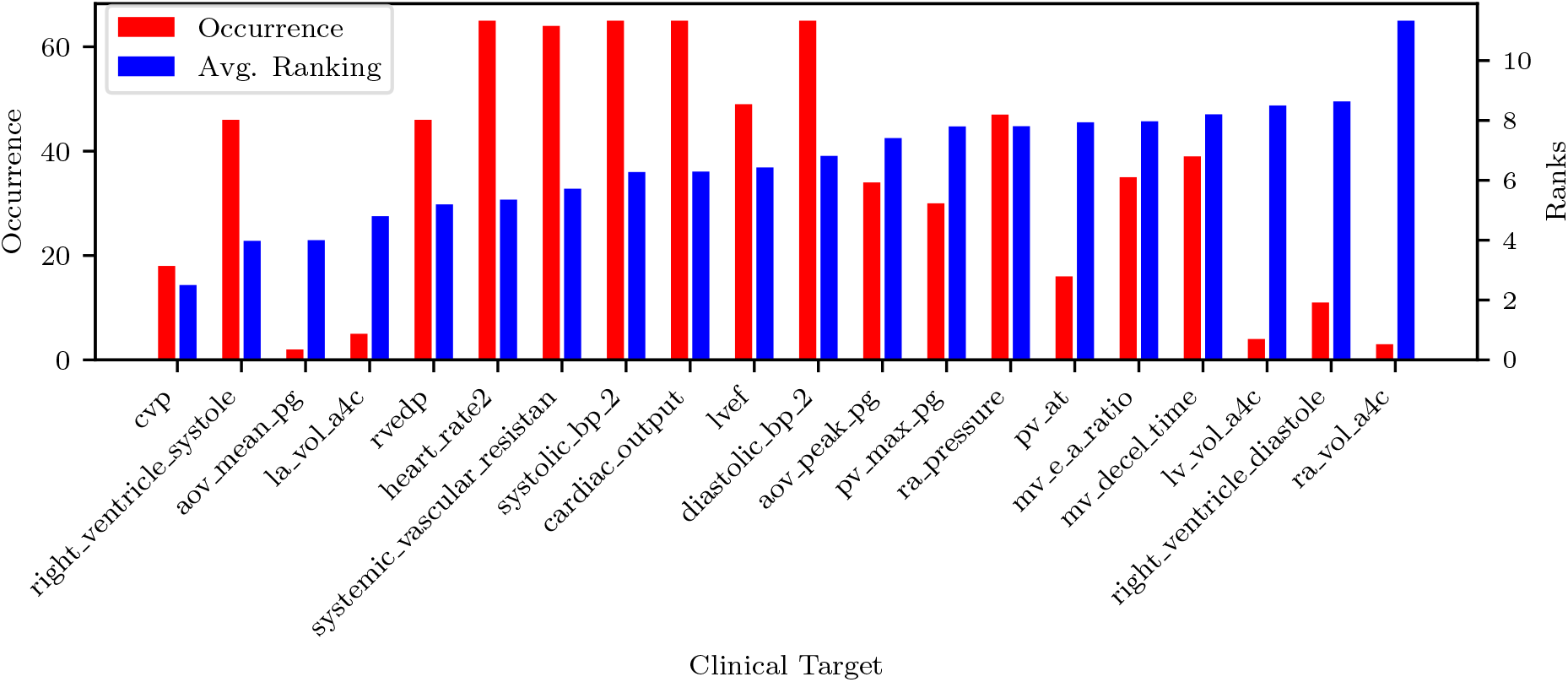
Target ranking and occurrence.

The list of targets ordered by average rank and filtered by occurrence is also shown in Table 5. Most of the quantities (except MPAP, PCW, PVR and SVR) can be estimated non-invasively. From RAP (or CVP, RVEDP), SBP-DBP, HR and SVR it is possible to estimate the cardiac output. From CO, HR and LVEF it is possible to estimate LVEDV which is correlated with PCW. This might explain why PCW is always estimated better than sPAP and dPAP. The mitral valve deceleration time and velocity E/A ratio are indicators of diastolic LV function and therefore correlated with PAP. If PVR, CO and PCW are known, then it is possible to estimate MPAP which is correlated to sPAP and dPAP. Finally the parameter ranking is summarized for all patients in Figure 16.

**Table 5:**
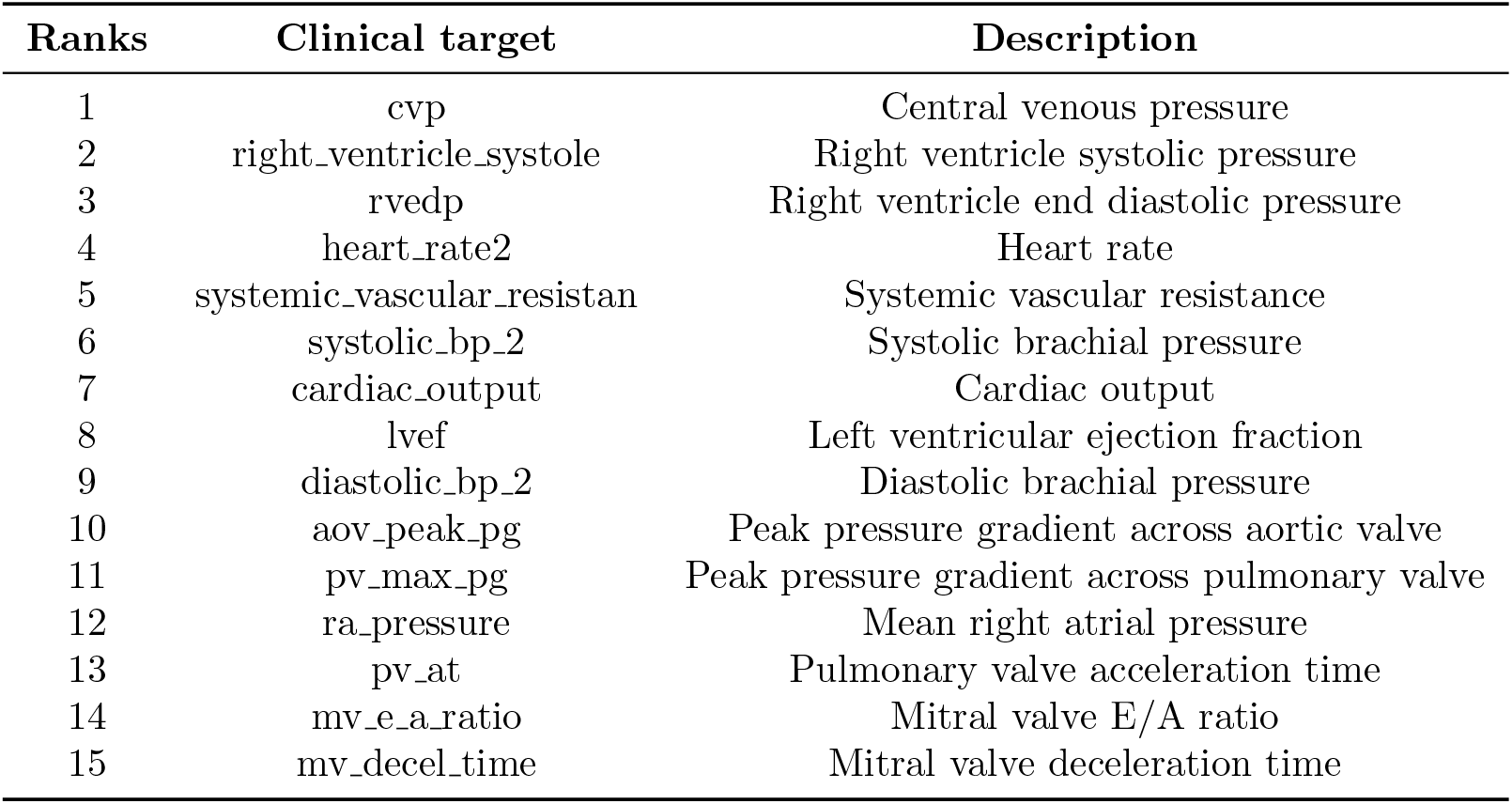
Target list in order of average rank, filtered by occurrence. Targets with less than 5 occurrences were excluded from the ranking order.

**Figure 16:**
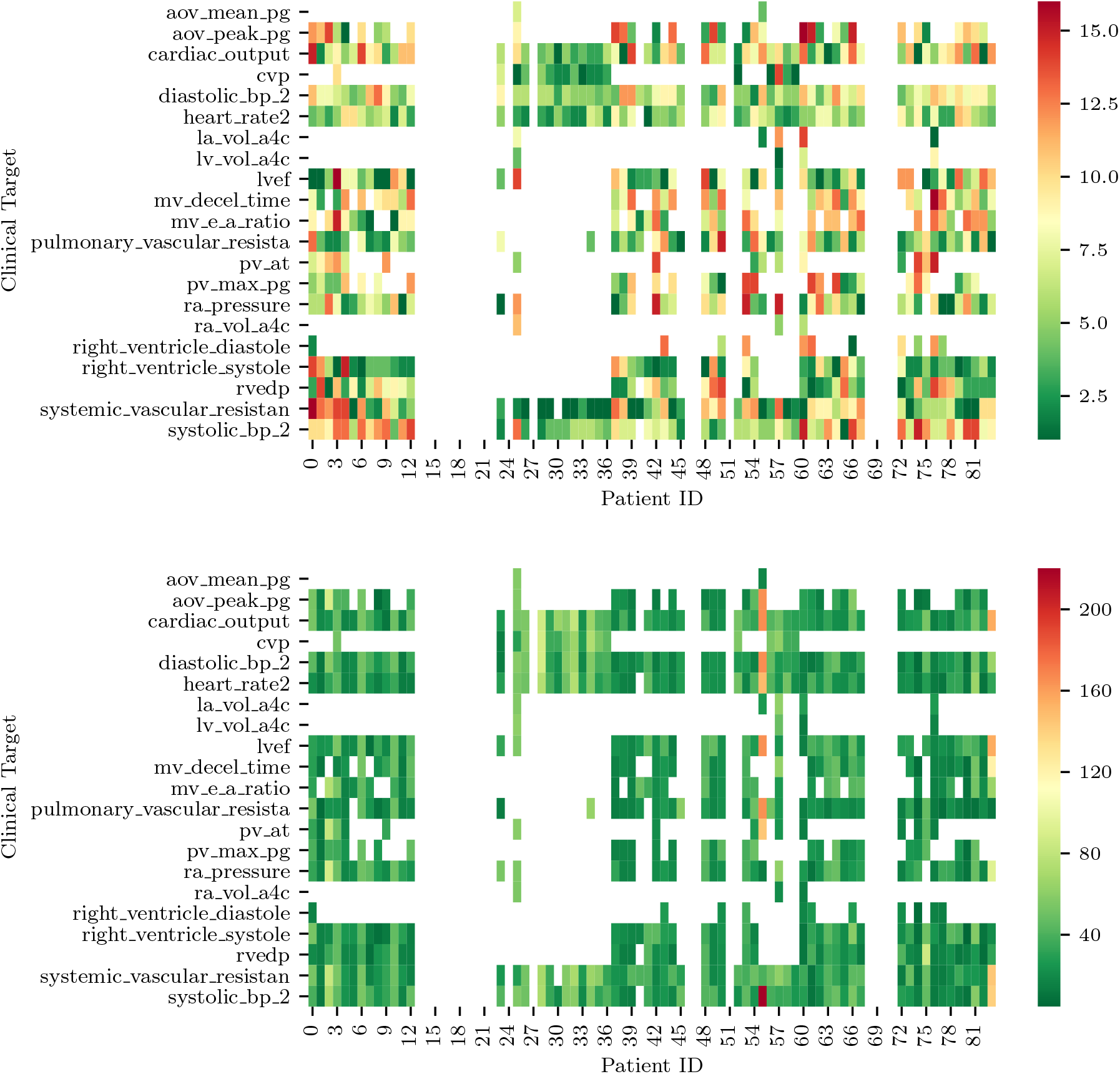
(a) Ranks of clinical targets for each patient in order of importance (i.e., green more importance, red less important). (b) Minimum combined prediction error associated with the introduction of each clinical target for all the patients included in the study. Note how blank entries correspond to missing clinical targets.

### 3.6 PH classifiers from assimilated circulation models

In this section, we explore the use of trained lumped parameter models for automatic detection of abnormal pulmonary pressures from minimally invasive clinical measurements. We employ a naïve Bayes classifiers [54] to detect pulmonary hypertension from our dataset. The first step of this classification process was to generate a ground truth variable for hypertension that could be used to analyze the accuracy of our hypertensive classification. Using the patients clinical data, a binary hypertensive variable was defined using the criteria from [43]: if a patient has mPAP*>*25 mmHG or sPAP*>*35 mmHG, then the patient is classified as hypertensive. Of the 82 patients, 65 of the patients contained sufficient clinical data to allow for the creation of a ground truth binary hypertensive variable. The remaining 17 patients did not contain enough clinical data to determine a ground truth value for hypertension, so these patients were excluded from the testing and training datasets.

Before a naïve Bayes classification can be performed, the problem of missing data must first be addressed. Our dataset representing the cohort of 82 patients contains a non-negligible ratio of missing data, with patients missing between 1 to 19 of the 24 total clinical targets. To overcome this issue and to verify how classification results depend on the strategy selected for missing data imputation, five different missing data imputation approaches were tested. The first method tested was *complete-case analysis*, considering only the 4 variables that were available for all patients, i.e., heart rate, systolic blood pressure, diastolic blood pressure, and cardiac output. The remaining four missing data methods consisted of replacing the missing values with (1) zeros, (2) the max value of the data set, (3) a value far outside the range of the data set (10 times the max), or (4) the median. In addition to the training of five separate classifiers using five different missing data methods, a sixth naïve Bayes classifier was trained on the MAP parameters of the model. The data for the MAP parameters from the model is a complete set, so no missing data method was required. Figure 17 shows how the above imputation approaches affect classification accuracy. At first sight, the high accuracy produced by a multiple imputation strategy using the maximum inter-patient clinical value (or 10 times its value) may seem surprising. This can be explained by observing how, in such a case, the probability of the feature associated with the missing data is essentially zero, thus essentially reducing the number of features and increasing the resulting accuracy.

**Figure 17:**
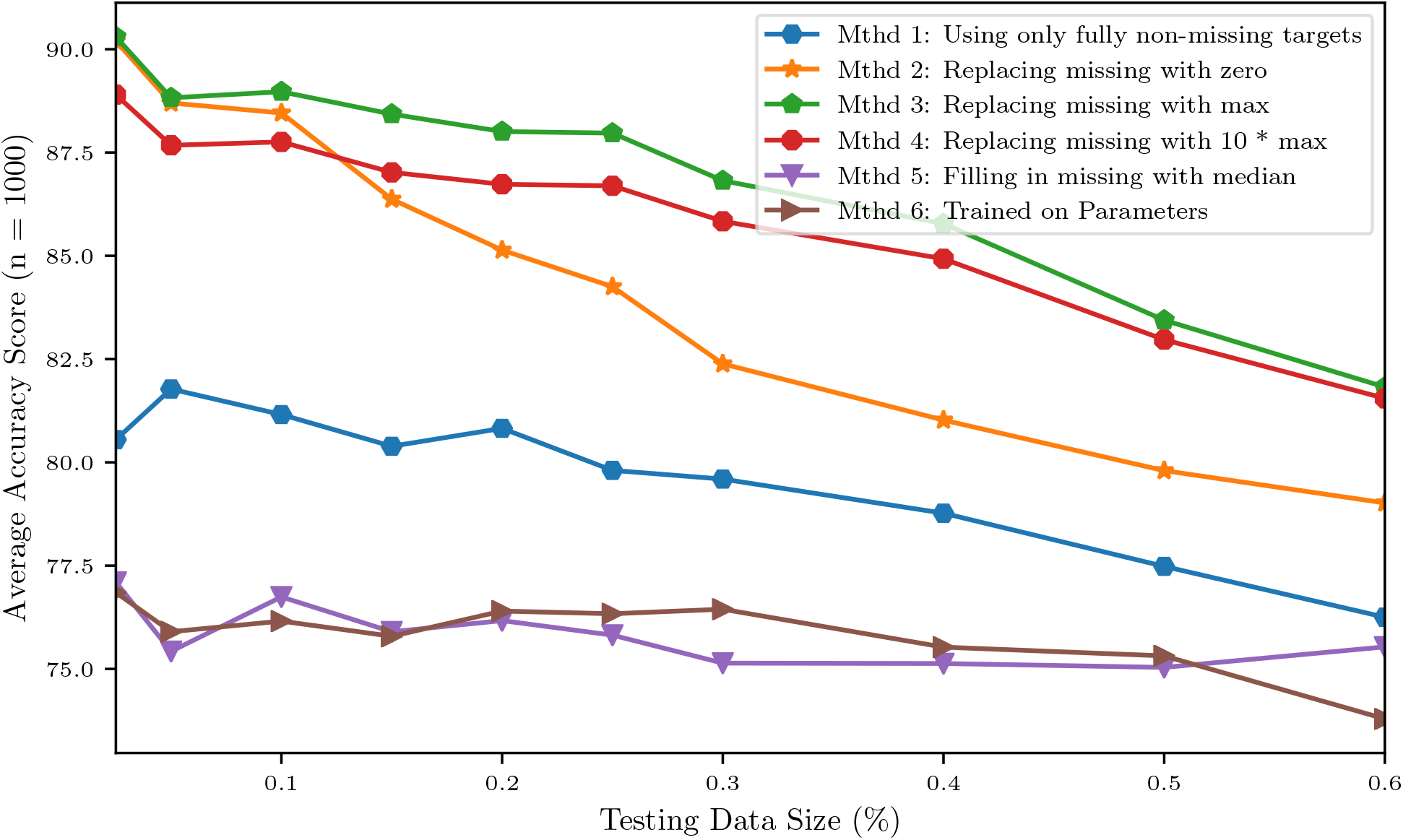
Accuracy of a naïve Bayes classifiers for pulmonary hypertension using different approaches for multiple imputation.

We then grouped patients according to a training and a testing set, consisting of 4*/*5 and 1*/*5 of the available data, respectively. Training and testing is performed either using the raw clinical data or model parameters assimilated though optimization and MCMC, resulting in approximately 88% accuracy for the classifier trained with the 24 raw clinical datapoints and about 76.4% accuracy for the classifier trained with the 45 assimilated parameter values (see Table 6).

**Table 6:** Training accuracy improvements using the original dataset and following pre-processing to balance the relatively small number of hypertensive positives.

|  |  | Pre-processing Data for Classification |  |  |
| --- | --- | --- | --- | --- |
|  |  | Complete Data | Missing Data Imputed | Balanced Data |
| Training data type for classifier | Clinical Data | 0.8082 | 0.8801 | 1.0 |
|  | Parameters | 0.7640 | N.A. | 1.0 |

Of the 82 patients, 65 contained enough information for ground truth classification of whether or not the patient should be identified as hypertensive. Based on these data we were able to classify 22 of the 65 patients, or about one third of the patients, as being affected by pulmonary hypertension and the remaining two thirds with normal PAP pressures. This imbalance is known to introduce bias [55], with the classifier more likely to label patients as non-hypertensive (as 2/3 of the training data are non-hypertensive). As a remedy, six different approaches were applied to deal with this unbalance, which represents a mixture of over-sampling methods, under-sampling methods, and a combination of both.

Figure 18 shows the principal component decomposition and contingency table for the unbalanced data and the data balanced with centroid clustering. Note how the confusion matrices are evaluated based on test data, hence 13 patients (1/5 of the 65 patients) for the unbalanced dataset. Undersampling in centroid clustering reduces the training/testing data from 65 to 33, hence one-fifth of the 33 samples becomes approximately 7, as shown in the confusion matrix for the centroid clustering under-sampling method.

**Figure 18:**
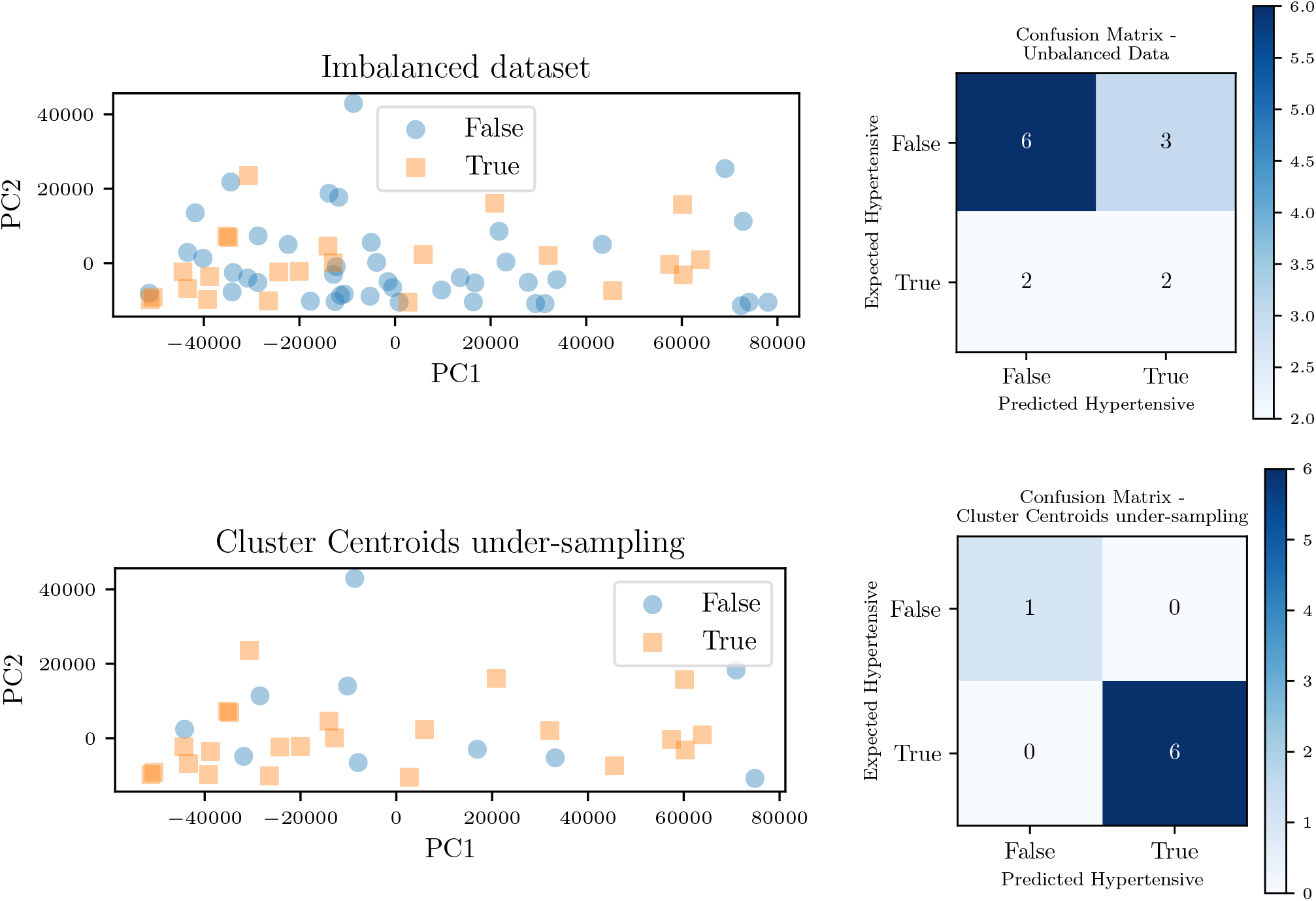
Principal component decomposition (left) along with the contingency table (right) for the unbalanced data and balanced through *centroid clustering*.

Figure 19 shows instead the resulting effect of each unbalanced data method on the overall area under the receiver operating characteristic curve (ROC) for the classifier trained on the model parameters. For such a classifier, centroid clustering [56] increases accuracy from 76.4% up to 100%. In addition, data balance using Synthetic Minority Oversampling TEchnique (SMOTE) [57] increases the accuracy of the model trained on the data from 88% to 100% as shown in Table 6. We again remark that the number of patients in this study is small and future studies will investigate generalization of this approach to larger datasets.

**Figure 19:**
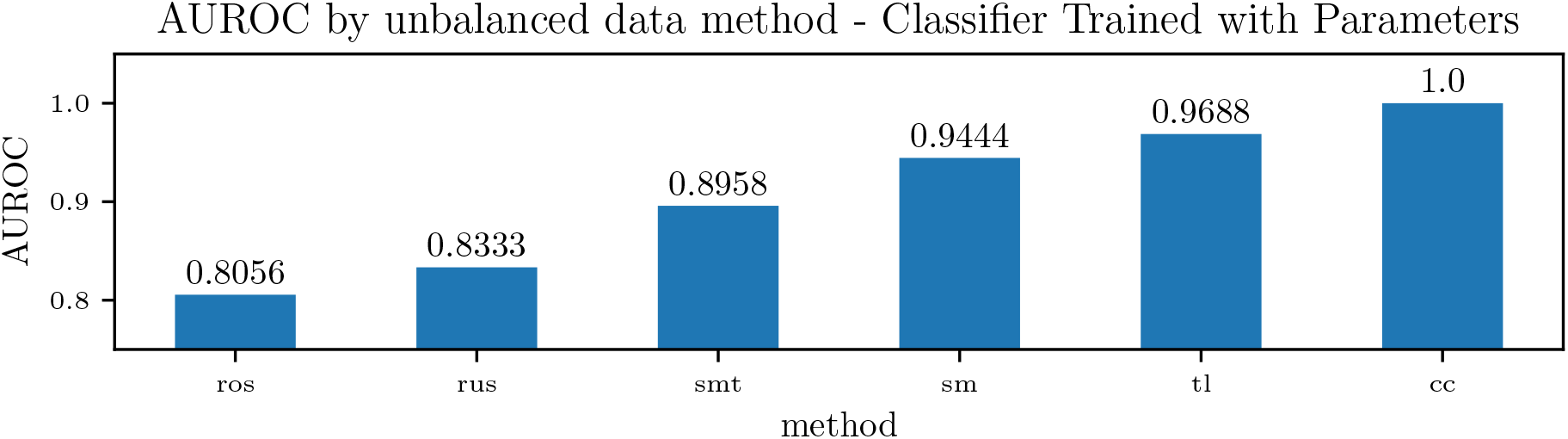
Accuracy of a Naïve Bayes classifiers for pulmonary hypertension, using identified lumped parameters as features, and various methods to handle unbalanced data. The methods used were Random Over-sampling (ros), Random Under-sampling (rus), an over-sampling method: SMOTE (Synthetic Minority Oversampling TEchnique) (sm), two under-sampling methods: Tomek Links (tl) and Cluster Centroids (cc), and a combination method that performs over-sampling followed by under-sampling through applying SMOTE followed by Tomek Links (smt).

## 4 Conclusion

This study demonstrates that a relatively simple lumped parameter compartmental model can represent a wide range of physiologies, spanning healthy patients to patients affected by severe diastolic left ventricular dysfunction. In this context, an activation formulation for the heart chambers has proven important to separately account for systolic and diastolic pressure-volume behavior. When trained using ideal clinical data from subjects with diastolic left ventricular dysfunction, the parameters associated with the governing physiological mechanisms (i.e., diastolic ventricle relaxation) change in a way that is consistent with the underlying physiology of the dysfunction. The average parameter sensitivities are determined after training with real data from 82 patients, confirming intuition for the most important parameters. Pulmonary pressures were found to be mostly sensitive to heart rate and contraction timing parameters, diastolic and systolic pressure/volume ratio parameters (*K*_*pas*,1,*lv*_, *K*_*pas*,2,*lv*_ and *E*_*max,lv*_), left atrioventricular and aortic valve resistance (*R*_*la,lv*_, *R*_*lv,ao*_), pulmonary resistance and capacitance (*R*_*pa*_, *C*_*pa*_) and systemic resistance (*R*_*sys*_). Additionally, the model was found to be locally unidentifiable, with initial conditions generally unimportant. The structural identifiability analysis showed that inference of model parameters is feasible under perfect, noiseless conditions and with a sufficiently large number of available clinical targets. Target ranking based on sequential optimization reveals the most important non-invasively acquired clinical targets to be the heart rate, systemic pressures, peak pressure gradient across aortic and pulmonary valves, pulmonary valve acceleration time, mitral valve deceleration time and mitral valve E/A peak ratio. Clinical targets requiring invasive measurement practices that positively affect the prediction of pulmonary pressures are instead central venous pressure, right ventricular systolic and diastolic pressure, systemic vascular resistance, cardiac output and mean right atrial pressure. After investigating parameter identifiability/sensitivity, pulmonary pressures in 82 patients with various heart failure severities were predicted from a lumped hemodynamic model, trained based on the remaining clinical targets. The average absolute pressure error on the 11 patients characterized by at least 11 distinct clinical entries was found approximately equal to 8 and 6 mmHg for systolic and diastolic/wedge pulmonary pressures, respectively. While these errors may seem large to detect cases of mild hypertension, they may be more than reasonable depending on how high are the PA pressures we are trying to predict/detect. In other words, while a 8 mmHg pressure error may seem relevant with an actual pulmonary pressure of 20 mmHg, for cases of patients with *real* disease (i.e. PA pressures of 40s, 50s, 60s, etc.) then the difference between (say) 50 and 58 would not be nearly as large, but more importantly our approach would still identify those patients requiring further evaluation.

Finally, we have shown that MAP estimates of circulation model parameters can be used to detect elevated pulmonary pressures, and that a simple classifier provide high accuracy on balanced data, even when these parameters are identified without measuring systolic, diastolic, wedge pulmonary pressures and pulmonary vascular resistance. Future work will be devoted to the systematic demonstration of the proposed approach on larger patient data sets, opening new avenues for translational applications of model-based diagnostics, improving model formulations targeting specific diseases and for the development of improved and more efficient estimation approaches.

## Data Availability

The EHR data set used in the paper will be made available as additional material when the paper will be published.

## Acknowledgements

This work was supported by a Google ATAP grant, a National Institute of Health grant 1R01EB018302, a National Science Foundation grant CDS&E (PI Alison Marsden) and a NSF Big Data Science & Engineering grant #1918692 (PI Daniele Schiavazzi). The authors would also like to thank Derek DeBusschere and Sandy Ng for their guidance and supervision. This research used computational resources provided through the Center for Research Computing at the University of Notre Dame.

## Conflict of Interest Statement

The authors declare that there is no conflict of interest.

## Appendices

### Appendix A Uniform priors and default parameter set

Table A1 shows the maximum and minimum ranges for each lumped parameter. The default parameter values used for the structural identifiability analysis in Section 3.3.2 are also reported.

**Table A1:** Maximum and minimum values defining the priors for the model parameters.

| Parameter | Symbol | Default<sup>(*)</sup> | Min<sup>(*)</sup> | Max<sup>(*)</sup> |
| --- | --- | --- | --- | --- |
| Heart Rate | HR | 78.0 | 40.0 | 100.0 |
| Atrial relative activation duration | $t_{sas}$ | 0.2 | 0.05 | 0.4 |
| Atrial relative activation time shift | $t_{pws}$ | 9.5 | 5.0 | 10.0 |
| Ventricular relative activation duration | $t_{svs}$ | 0.4 | 0.1 | 0.5 |
| Atrial passive curve slope, right atrium | $K_{pas,ra,1}$ | 5.0 | 0.1 | 10.0 |
| Atrial passive curve exponent factor, right atrium | $K_{pas,ra,2}$ | 0.006 | 0.0001 | 0.06 |
| Atrial active curve slope, right atrium | $E_{max,ra}$ | 0.1 | 0.05 | 5.0 |
| Unstressed right atrial volume | $V_{ra,0}$ | 0.0 | 0.0 | 50.0 |
| Atrial passive curve slope, left atrium | $K_{pas,la,1}$ | 5.0 | 0.1 | 10.0 |
| Atrial passive curve exponent factor, left atrium | $K_{pas,la,2}$ | 0.0065 | 0.0001 | 0.06 |
| Atrial active curve slope, left atrium | $E_{max,la}$ | 0.2 | 0.05 | 5.0 |
| Unstressed left atrial volume | $V_{la,0}$ | 0.0 | 0.0 | 50.0 |
| Ventricular passive curve slope, right ventricle | $K_{pas,rv,1}$ | 5.0 | 0.1 | 20.0 |
| Ventricular passive curve exponent factor, right ventricle | $K_{pas,rv,2}$ | 0.03 | 0.0001 | 0.01 |
| Ventricular active curve slope, right ventricle | $E_{max,rv}$ | 0.5 | 0.1 | 5.0 |
| Unstressed right atrial volume | $V_{rv,0}$ | 0.0 | 0.0 | 50.0 |
| Ventricular passive curve slope, left ventricle | $K_{pas,lv,1}$ | 2.0 | 0.1 | 20.0 |
| Ventricular passive curve exponent factor, left ventricle | $K_{pas,lv,2}$ | 0.003 | 0.0001 | 0.01 |
| Ventricular active curve slope, left ventricle | $E_{max,lv}$ | 4.0 | 1.0 | 5.0 |
| Unstressed left atrial volume | $V_{lv,0}$ | 20.0 | 0.0 | 50.0 |
| Inductance of right atrium | $L_{ra,rv}$ | 0.1 | 0.1 | 0.1 |
| Resistance of right atrium | $R_{ra,rv}$ | 10.0 | 10.0 | 10.0 |
| Inductance of right ventricle | $L_{rv,pa}$ | 0.1 | 0.1 | 0.1 |
| Resistance of right ventricle | $R_{rv,pa}$ | 15.0 | 15.0 | 15.0 |
| Inductance of left atrium | $L_{la,lv}$ | 0.1 | 0.1 | 0.1 |
| Resistance of left atrium | $R_{la,lv}$ | 8.0 | 8.0 | 8.0 |
| Inductance of left ventricle | $L_{lv,ao}$ | 0.1 | 0.1 | 0.1 |
| Resistance of left ventricle | $R_{lv,ao}$ | 25.0 | 25.0 | 25.0 |
| Aortic capacitance | $C_{ao}$ | 1.0e-3 | 1.0e-5 | 0.001 |
| Pulmonary capacitance | $C_{pa}$ | 4.0e-3 | 100.0e-6 | 0.01 |
| Pulmonary resistance | $R_{pa}$ | 130.0 | 1.0 | 500.0 |
| Systemic capacitance | $C_{sys}$ | 400.0e-6 | 100.0e-6 | 0.05 |
| Systemic Resistance - Arteries | $R_{sys,a}$ | 400.0 | 100.0 | 800.0 |
| Systemic Resistance - Veins | $R_{sys,v}$ | 1200.0 | 500.0 | 2500.0 |

(\*)All units are in the CGS system.

## Notes

### Competing Interest Statement

The authors have declared no competing interest.

### Author Declarations

Compliance with Ethical Standards - The study was classified as research not involving human subjects and approved on June 13th, 2019 by the Office of Research Compliance and Institutional Review Board at the University of Notre Dame under IRB#19-05-5371. All procedures performed in studies involving human participants were in accordance with the ethical standards of the institutional and/or national research committee and with the 1964 Helsinki declaration and its later amendments or comparable ethical standards. For this retrospective study formal consent is not required.

## References

[1] M. Obokata, Y.N.V. Reddy, and B.A. Borlaug. Diastolic dysfunction and heart failure with preserved ejection fraction: understanding mechanisms by using noninvasive methods. JACC: Cardiovascular Imaging, 13(1):245–257, 2020.

[2] R.O. Bonow and J.E. Udelson. Left ventricular diastolic dysfunction as a cause of congestive heart failure: mechanisms and management. Annals of Internal Medicine, 117(6):502–510, 1992.

[3] D. Mozaffarian, E.J. Benjamin, A.S. Go, D.K. Arnett, M.J. Blaha, M. Cushman, S.R. Das, S. de Ferranti, J-P. Després, H.J. Fullerton, et al. Executive summary: Heart disease and stroke statistics-2016 update: A report from the american heart association. Circulation, 133(4):447, 2016.

[4] M.R. Zile and D.L. Brutsaert. New concepts in diastolic dysfunction and diastolic heart failure: Part i diagnosis, prognosis, and measurements of diastolic function. Circulation, 105(11):1387–1393, 2002.

[5] C.S.P. Lam, V.L. Roger, R.J. Rodeheffer, B.A. Borlaug, F.T. Enders, and M.M. Redfield. Pulmonary hypertension in heart failure with preserved ejection fraction: a community-based study. Journal of the American College of Cardiology, 53(13):1119–1126, 2009.

[6] S.J Shah, D.H. Katz, S. Selvaraj, M.A. Burke, C.W. Yancy, M. Gheorghiade, R.O. Bonow, C-C. Huang, and R.C. Deo. Phenomapping for novel classification of heart failure with preserved ejection fraction. Circulation, 131(3):269–279, 2015.

[7] M.R. Fisher, P.R. Forfia, E. Chamera, T. Housten-Harris, H.C. Champion, R.E. Girgis, M.C. Corretti, and P.M. Hassoun. Accuracy of doppler echocardiography in the hemodynamic assessment of pulmonary hypertension. American Journal of Respiratory and Critical Care Medicine, 179(7):615–621, 2009.

[8] N. Galiè, M.M. Hoeper, M. Humbert, A. Torbicki, J.L. Vachiery, J.A. Barbera, M. Beghetti, P. Corris, S. Gaine, J.S. Gibbs, et al. Guidelines for the diagnosis and treatment of pulmonary hypertension. European Heart Journal, 30(20):2493–2537, 2009.

[9] M.F. Snyder and V.C. Rideout. Computer simulation studies of the venous circulation. IEEE Transactions on Biomedical Engineering, BME–16(4):325–334, 1969.

[10] G. Avanzolini, P. Barbini, A. Cappello, and A. Cevese. Time-varying mechanical properties of the left ventricle-a computer simulation. IEEE Transactions on Biomedical Engineering, BME-32(10):756– 763, 1985.

[11] G. Avanzolini, P. Barbini, A. Cappello, and G. Cevenini. CADCS simulation of the closed-loop cardiovascular system. International journal of bio-medical computing, 22(1):39–49, 1988.

[12] M. Ursino. Interaction between carotid baroregulation and the pulsating heart: a mathematical model. American Journal of Physiology-Heart and Circulatory Physiology, 275(5):H1733–H1747, 1998.

[13] G. Pennati, F. Migliavacca, G. Dubini, R. Pietrabissa, and M.R. de Leval. A mathematical model of circulation in the presence of the bidirectional cavopulmonary anastomosis in children with a univentricular heart. Medical engineering & physics, 19(3):223–234, 1997.

[14] Francesco Migliavacca, Giancarlo Pennati, Gabriele Dubini, Roberto Fumero, Riccardo Pietrabissa, Gonzalo Urcelay, Edward L. Bove, Tain-Yen Hsia, and Marc R. de Leval. Modeling of the norwood circulation: effects of shunt size, vascular resistances, and heart rate. American Journal of Physiology-Heart and Circulatory Physiology, 280(5):H2076–H2086, 2001.

[15] G. Pennati, F. Migliavacca, G. Dubini, and E.L. Bove. Modeling of systemic-to-pulmonary shunts in newborns with a univentricular circulation: state of the art and future directions. Progress in Pediatric Cardiology, 30(1-2):23–29, 2010.

[16] B.A. Deswysen. Parameter estimation of a simple model of the left ventricle and of the systemic vascular bed, with particular attention to the physical meaning of the left ventricular parameters. IEEE Transactions on Biomedical Engineering, (1):29–38, 1977.

[17] B. Deswysen, A.A. Charlier, and M. Gevers. Quantitative evaluation of the systemic arterial bed by parameter estimation of a simple model. Medical and Biological Engineering and Computing, 18(2):153–166, 1980.

[18] J.W. Clark, R.Y.S. Ling, R. Srinivasan, J.S. Cole, and R.C. Pruett. A two-stage identification scheme for the determination of the parameters of a model of left heart and systemic circulation. IEEE Transactions on Biomedical Engineering, (1):20–29, 1980.

[19] B. McInnis, Z-W. Guo, P. Lu, and J-C. Wang. Adaptive control of left ventricular bypass assist devices. IEEE Transactions on automatic control, 30(4):322–329, 1985.

[20] T. Shimooka, Y. Mitamura, and T. Yuhta. Investigation of parameter estimator and adaptive controller for assist pump by computer simulation. Artificial organs, 15(2):119–128, 1991.

[21] Y-C. Yu, J.R. Boston, M.A. Simaan, and J.F. Antaki. Estimation of systemic vascular bed parameters for artificial heart control. IEEE Transactions on Automatic Control, 43(6):765–778, 1998.

[22] Y.C. Yu, J.R. Boston, M.A. Simaan, and J.F. Antaki. Minimally invasive estimation of systemic vascular parameters. Annals of biomedical engineering, 29(7):595–606, 2001.

[23] T.L. Ruchti, R.H. Brown, D.C. Jeutter, and X. Feng. Identification algorithm for systemic arterial parameters with application to total artificial heart control. Annals of biomedical engineering, 21(3):221–236, 1993.

[24] J.A. Revie, D.J. Stevenson, J.G. Chase, C.E. Hann, B.C. Lambermont, A. Ghuysen, P. Kolh, G.M. Shaw, S. Heldmann, and T. Desaive. Validation of subject-specific cardiovascular system models from porcine measurements. Computer methods and programs in biomedicine, 109(2):197–210, 2013.

[25] K. Sughimoto, F. Liang, Y. Takahara, K. Mogi, K. Yamazaki, S. Takagi, and H. Liu. Assessment of cardiovascular function by combining clinical data with a computational model of the cardiovascular system. The Journal of thoracic and cardiovascular surgery, 145(5):1367–1372, 2013.

[26] R.L. Spilker and C.A. Taylor. Tuning multidomain hemodynamic simulations to match physiological measurements. Annals of biomedical engineering, 38(8):2635–2648, 2010.

[27] A. Cappello, G. Cevenini, and G. Avanzolini. Model selection for ventricular mechanics: a sensitivity analysis approach. Journal of biomedical engineering, 9(1):13–20, 1987.

[28] G. Avanzolini, P. Barbini, and A. Cappello. Comparison of algorithms for tracking short-term changes in arterial circulation parameters. IEEE transactions on biomedical engineering, 39(8):861–867, 1992.

[29] D.E. Schiavazzi, A. Baretta, G. Pennati, T.Y. Hsia, and A.L. Marsden. Patient-specific parameter estimation in single-ventricle lumped circulation models under uncertainty. International journal for numerical methods in biomedical engineering, 33(3):e02799, 2017.

[30] J.S. Tran, D.E. Schiavazzi, A.B. Ramachandra, A.M. Kahn, and A.L. Marsden. Automated tuning for parameter identification and uncertainty quantification in multi-scale coronary simulations. Computers & fluids, 142:128–138, 2017.

[31] C.M. Witzenburg and J.W. Holmes. Predicting the time course of ventricular dilation and thickening using a rapid compartmental model. Journal of cardiovascular translational research, 11(2):109–122, 2018.

[32] C. Luo, D. Ramachandran, D.L. Ware, T.S. Ma, and J.W. Clark. Modeling left ventricular diastolic dysfunction: classification and key indicators. Theoretical Biology and Medical Modelling, 8(1):1, 2011.

[33] S. Tewari, S. Bugenhagen, Z. Wang, D. Schreier, B. Carlson, N. Chesler, and D. Beard. Analysis of cardiovascular dynamics in pulmonary hypertensive C57BL6/J mice. Frontiers in physiology, 4:355, 2013.

[34] P. Del Moral, A. Doucet, and A. Jasra. Sequential monte carlo samplers. Journal of the Royal Statistical Society: Series B (Statistical Methodology), 68(3):411–436, 2006.

[35] Y. LeCun, Y. Bengio, and G. Hinton. Deep learning. nature, 521(7553):436–444, 2015.

[36] J.L. Schafer. Multiple imputation: a primer. Statistical methods in medical research, 8(1):3–15, 1999.

[37] V.C. Rideout and D.E. Dick. Difference-differential equations for fluid flow in distensible tubes. IEEE Transactions on Biomedical Engineering, (3):171–177, 1967.

[38] S. Pant, C. Corsini, C. Baker, T.Y. Hsia, G. Pennati, I.E. Vignon-Clementel, and Modeling of Congenital Hearts Alliance (MOCHA) Investigators. Data assimilation and modelling of patient-specific single-ventricle physiology with and without valve regurgitation. Journal of biomechanics, 49(11):2162–2173, 2016.

[39] Edwards Lifesciences Corporation. Normal hemodynamic parameters and laboratory values. http://ht.edwards.com/scin/edwards/sitecollectionimages/edwards/products/pre-sep/ar04313hemodynpocketcard.pdf.

[40] K. Yared, P. Noseworthy, A.E. Weyman, E. McCabe, M.H. Picard, and A.L. Baggish. Pulmonary artery acceleration time provides an accurate estimate of systolic pulmonary arterial pressure during transthoracic echocardiography. Journal of the American Society of Echocardiography, 24(6):687–692, 2011.

[41] Elaine P Gordon, Ingela Schnittger, Peter J Fitzgerald, Paul Williams, and Richard L Popp. Reproducibility of left ventricular volumes by two-dimensional echocardiography. Journal of the American College of Cardiology, 2(3):506–513, 1983.

[42] AM Maceira, SK Prasad, M Khan, and DJ Pennell. Normalized left ventricular systolic and diastolic function by steady state free precession cardiovascular magnetic resonance. Journal of Cardiovascular Magnetic Resonance, 8(3):417–426, 2006.

[43] G. Simonneau, M.A. Gatzoulis, I. Adatia, C. Celermajer, D.and Denton, A. Ghofrani, M.A. Sanchez Gomez, R.K. Kumar, M. Landzberg, R.F. Machado, H. Olschewski, I.M. Robbins, and Souza R. Updated clinical classification of pulmonary hypertension. Journal of the American College of Cardiology, 62(25 Supplement):D34–D41, 2013.

[44] T.M. Cover. Geometrical and statistical properties of systems of linear inequalities with applications in pattern recognition. IEEE Transactions on Electronic Computers, EC–14(3):326–334, 1965.

[45] John A Nelder and Roger Mead. A simplex method for function minimization. The computer journal, 7(4):308–313, 1965.

[46] J.A. Vrugt, C.J.F. Ter Braak, C.G.H. Diks, B.A. Robinson, J.M. Hyman, and D. Higdon. Accelerating Markov chain Monte Carlo simulation by differential evolution with self-adaptive randomized subspace sampling. International Journal of Nonlinear Sciences and Numerical Simulation, 10(3):273–290, 2009.

[47] J.A. Vrugt. Markov chain Monte Carlo simulation using the DREAM software package: Theory, concepts, and MATLAB implementation. Environmental Modelling & Software, 75:273–316, 2016.

[48] A. Gelman and D.B. Rubin. Inference from iterative simulation using multiple sequences. Statistical Science, pages 457–472, 1992.

[49] D.E. Schiavazzi, A. Baretta, G. Pennati, T.Y. Hsia, and A.L. Marsden. Patient-specific parameter estimation in single-ventricle lumped circulation models under uncertainty. International Journal for Numerical Methods in Biomedical Engineering, 2016. In Press.

[50] A.R. Akintunde, K.S. Miller, and D.E. Schiavazzi. Bayesian inference of constitutive model parameters from uncertain uniaxial experiments on murine tendons. Journal of the mechanical behavior of biomedical materials, 96:285–300, 2019.

[51] S. Kullback and R.A. Leibler. On information and sufficiency. The annals of mathematical statistics, 22(1):79–86, 1951.

[52] G.P. Tracy, M.S. Proctor, and C.S. Hizny. Reversibility of pulmonary artery hypertension in aortic stenosis after aortic valve replacement. The Annals of thoracic surgery, 50(1):89–93, 1990.

[53] T.J. Rothenberg. Identification in parametric models. Econometrica: Journal of the Econometric Society, pages 577–591, 1971.

[54] D.D. Lewis. Naive (Bayes) at forty: The independence assumption in information retrieval. In European Conference on Machine Learning, pages 4–15. Springer, 1998.

[55] Y. Sun, A.K.C. Wong, and M.S. Kamel. Classification of imbalanced data: a review. International Journal of Pattern Recognition and Artificial Intelligence, 23(04):687–719, 2009.

[56] W.C. Lin, C.F. Tsai, Y.H. Hu, and J.S. Jhang. Clustering-based undersampling in class-imbalanced data. Information Sciences, 409:17–26, 2017.

[57] N.V. Chawla, K.W. Bowyer, L.O. Hall, and W.P. Kegelmeyer. Smote: synthetic minority over- sampling technique. Journal of artificial intelligence research, 16:321–357, 2002.

